# Epigenome-wide association study of cerebrospinal fluid-based biomarkers of Alzheimer’s disease in cognitively normal individuals

**DOI:** 10.1101/2025.02.04.25321657

**Authors:** Anke Hüls, Jiaqi Liu, Chaini Konwar, Karen N. Conneely, Allan I. Levey, James J. Lah, Aliza P. Wingo, Thomas S. Wingo

## Abstract

**INTRODUCTION:** Cerebrospinal fluid (CSF) biomarkers of Alzheimer’s disease (AD) are reliable predictors of future AD risk. We investigated whether pre-clinical changes in AD CSF biomarkers are reflected in blood DNA methylation (DNAm) levels in cognitively normal participants.

**METHODS:** We profiled blood-based DNAm with the EPIC array in participants without a diagnosis of cognitive impairment in the Emory Healthy Brain Study (EHBS; N=495) and ADNI (N=122). Their CSF Aβ42, tTau, and pTau levels were quantified using Elecsys immunoassays. We conducted epigenome-wide association studies to assess associations between DNAm and CSF biomarkers of AD.

**RESULTS:** In EHBS, no loci were Bonferroni-significant after adjusting for confounding factors. In ADNI, two loci were significant, but they were not replicated in EHBS. There was little agreement between the top loci from EHBS and ADNI.

**DISCUSSION:** Our study showed little evidence of an association between differential blood-based DNAm and pre-clinical AD CSF biomarkers.

## Background

Alzheimer’s disease and related dementias (AD/ADRD) are the 6^th^ leading cause of death affecting over six million people in the US, and the number is projected to grow to 13.8 million by 2060 (1,2). Individuals living with AD/ADRD experience progressively worsening cognitive impairment that compromises their quality of life and often requiring long-term day-to-day support and care (2).

Alzheimer’s disease (AD) has a long asymptomatic stage that can be detected in brain imaging and cerebrospinal fluid (CSF) (3). In 2018, the National Institute on Aging and Alzheimer’s Association drew a distinction between the underlying pathologic process of AD and the clinical signs and symptoms those pathologic changes may cause (i.e., subjective cognitive impairment, mild cognitive impairment [MCI], and dementia). The rationale for this distinction is the emergence of reliable AD biomarkers that can measure beta-amyloid deposition, pathologic tau, and neurodegeneration (collectively termed the ATN framework), which are detectable up to 30 years *before* symptomatic cognitive impairment (4). Identifying patients who are on the path of developing dementia is essential for early intervention and treatment. Early biomarkers of AD, e.g., CSF biomarkers (5–9), provide an opportunity to detect the first signs of disease.

In recent years, there has been a substantial increase in studies characterizing epigenetic mechanisms in AD, mainly focused on DNA methylation (DNAm) (10). In the brain, epigenetic studies suggest robust associations between DNAm and AD (11), including differential DNAm in the amyloid precursor protein (*APP*) (12), microtubule-associated protein tau (*MAPT*) (12), apolipoprotein (*APOE*) promoter region (13), homeobox A3 (*HOXA3*) (14), interleukin-1 beta (*IL-1*β) (15), interleukin-6 (*IL-6*) (15), and claudin-5 (*CLDN5*) genes (16). While identifying AD-related epigenetic signatures in the brain can help unravel some biological mechanisms involved in the pathophysiology of AD, understanding epigenetic changes in the blood is crucial for the development of new molecular indicators for AD, given that blood collection is a minimal invasive routine procedure, which can be performed while patients are alive (17).

Several blood-based DNAm markers have been identified in association with AD, including differential DNAm in the homeobox B6 (*HOXB6*) gene in AD (18), the oxytocin (*OXT*) gene (19), the adenosine deaminase RNA–specific B2 (*ADARB2*) gene (20). One study of 202 participants from the Alzheimer’s Disease Neuroimaging Initiative (ADNI) cohort identified a number of associations between blood DNAm and CSF biomarkers comparing 123 cognitively normal participants to 79 AD participants (21). Another study identified 12 DNAm sites in blood associated with CSF biomarkers among 885 participants included in the European Medical Information Framework for AD (EMIF-AD) study (22) – most of them diagnosed with mild cognitive impairment or AD. However, it is unknown whether differential DNAm is also associated with pre-clinical stages of AD.

To address this knowledge gap, we investigated whether blood DNAm was associated with CSF Aβ42, phosphorylated tau (pTau), and total tau (tTau) biomarkers in 617 cognitively normal participants enrolled in the Emory Healthy Brain Study (EHBS) and the ADNI cohort. We hypothesized that pre-clinical changes in CSF biomarkers of AD will be mirrored in the blood epigenome, which could help us to better understand the biological mechanisms underlying pre-clinical changes of AD. To test this hypothesis, we conducted independent epigenome-wide association studies in each cohort followed by an epigenome-wide meta-analysis to assess the association between blood-based DNAm and CSF biomarkers of AD.

## Methods

### Study population

Our study is based on data from 495 individuals from the EHBS (N=450 White participants, N=45 Black participants) and 122 White individuals from ADNI classified as controls without a diagnosis of cognitive impairment (i.e., mild cognitive impairment, Alzheimer’s disease or other dementias) at blood draw. Only one ADNI participant self-identified as Black and was excluded from the analysis. For EHBS, study inclusion criteria were availability of AD CSF biomarkers and blood to derive DNAm measures among the study participants in 2019.

The EHBS is a prospective research study focusing on preclinical biomarkers and the cognitive health of older adults. The EHBS is nested within the Emory Healthy Aging Study (EHAS) and includes participants from the metro-Atlanta region in Georgia, USA. The EHBS was launched in 2016 and the primary aim is to characterize biological, psychological and psychosocial factors associated with normal and abnormal aging through assessment of the central nervous system among adults 45-75 years old who were free of cognitive impairment in addition to several other chronic conditions (e.g. congestive heart failure, multiple sclerosis, HIV) at enrollment; more details on recruitment and eligibility have been published elsewhere(23). All participants completed an online consent process prior to enrollment and provided informed consent. The study was approved by the Emory University Institutional Review Board (IRB).

ADNI is a longitudinal, observational study with participant ages ranging from 55 to 90 designed to collect and validate biomarkers for AD. ADNI was launched in 2003 as a public-private partnership with a primary goal to test whether serial magnetic resonance imaging (MRI), positron emission tomography (PET), other biological markers, and clinical and neuropsychological assessment can be combined to improve measure the progression of mild cognitive impairment (MCI) and AD for clinical trials. Participant recruitment for ADNI is approved by the Institutional Review Board of each participating site. All ADNI participants undergo standardized diagnostic assessment that renders a clinical diagnosis of either control, MCI, or AD using standard research criteria (24).

### CSF AD biomarker measurements

In the EHBS, CSF biospecimens were collected via lumbar puncture at enrollment as previously described (25). Aβ42, tTau, and pTau CSF levels were quantified using the ElectroChemiLuminescense Immunoassay (ECLIA) Elecsys® AD CSF portfolio on an automated Roche Diagnostics instrument (F. Hoffman-La Roche Ltd). In ADNI, Aβ42, tTau, and pTau CSF levels were quantified using the Elecsys immunoassay detection platform (Roche Diagnostics Corporation, Indianapolis, IN USA) (26).

### DNA methylation data quality control

In EHBS and ADNI, DNAm was measured using the Illumina Infinium HumanMethylationEPIC BeadChip version 1, which quantified more than 850,000 CpGs sites in March 2020. DNAm preprocessing and sample quality control followed prior work (27,28) as described previously (29) and was done separately for EHBS and ADNI. Sample quality control measures included assessment of : a) 17 technical parameters with R package ewastools (30), including array staining, extension, hybridization, target removal, specificity, bisulfite conversion; b) estimate the methylated and unmethylated intensities with the getQC function from minfi (31) using default parameters; c) agreement between predicted (inferred from DNAm intensities of the sex chromosome using the minfi R package) and recorded sex; d) poor probe detection determined by if a samples had >1% probes with detection p-value > 0.01; e) low beadcount if 1% probes had beadcount < 3; f) outliers detection by function outlyx from R package wateRmelon(32) using default parameter.

After sample quality checks, probe level quality control was performed by removing XY probes and removing probes with bad detection p-value (p-value > 0.01) or with bead count < 3. Probes identified as poorly performing in more than 1% samples were removed from all samples. Probes that were cross hybridizing/cross reactive and occurred over polymorphic sites, defined by Pidsley annotation(33), were removed. After quality control, 661,869 probes and 450 unique samples remained for the analysis in the EHBS and 699,218 probes and 122 unique samples in ADNI.

Subsequently, probe-level normalization was done in two steps. Firstly, we normalized for color bias, background noise and dye-bias as implemented with preprocessNoob function from the minfi R package 1.42.0(34). Secondly, we applied the β-mixture quantile normalization (BMIQ) procedure in the wateRmelon R package 2.2.0(32) to normalize beta values distributions of type 1 and type 2 design probes in the Illumina arrays.

To account for batch effects, including chip ID, chip position, and plate effects, we used the function ComBat() from the sva R package 3.48.0(32) using default parameters.

### Epigenome-wide association study

Since the White EHBS participants (N=450) were by far the largest sample and CSF AD biomarkers differ by race (35), the White EHBS samples were used in our main analysis, followed by a replication of the top 10 CpGs in ADNI (N=122) and in the Black EHBS participants (N=45). We conducted independent epigenome-wide association studies (EWAS) of CSF AD biomarkers in EHBS (stratified by race) and ADNI (all White). We further conducted a replication analysis of the top 10 CpGs from ADNI in the White EHBS participants. An epigenome-wide meta-analysis of the White participants from EHBS and ADNI was conducted as a secondary analysis.

The main outcomes considered were tTau, pTau and Aβ42/Tau ratio, which were Box-Cox transformed using the *car* R package to improve normality. We also used ADNI-established thresholds (36) of CSF Aβ42 and pTau to dichotomize individuals for each measure (i.e., Aβ42+/- and pTau+/-). The threshold for Aβ42 was 980 pg/ml, and threshold for pTau was 21.8 pg/ml. We used ADNI-established thresholds of CSF Aβ_42_ < 980 pg/ml and pTau_181_ > 21.8 pg/ml to categorize individuals as either positive or negative for the respective measure (A+T+, A-T+, A+T-, and A-T-). The thresholds were selected to maximize the concordance with positive amyloid-β as determined by positive florbetapir (^18^F-AV-45) positron emission tomography (PET) imaging (26).

The association between DNAm beta values, which represent the ratio of the methylated probe intensity to the overall intensity (sum of methylated and unmethylated probe intensities) at a given CpG site, and CSF biomarkers (i.e., tTau, pTau, Aβ42/Tau ratio, Aβ42+/-, pTau+/-) was assessed using robust linear regression models with CSF biomarkers as independent variable and DNAm beta values as the dependent variable, with models fit using ’rlm’ from the MASS R package. All models were adjusted for age at baseline, sex, smoking status (i.e., with or without smoking history), and estimated cell-type proportions (i.e., B lymphocytes, natural killer cells, CD4□+□T lymphocytes, CD8□+□T lymphocytes, monocytes, and neutrophils). Cell type proportions were estimated using the ’FlowSorted.Blood.EPIC’ (version 1.8.0) R package 2.0.0 (37). To meta-analyze individual CpG results from EHBS and ADNI, we used the inverse-variance weighted fixed-effects model, as implemented in METAL(38). We used the Bonferroni threshold to account for multiple testing, resulting in a threshold of *p* = 7.55 x 10^-8^ or (0.05/66,1869) for the EWAS in EHBS and *p* = 7.60 x 10^-8^ (0.05/657,799) in the meta-analysis.

We also explored if we could find differentially methylated regions (DMRs) in relation to CSF AD biomarkers using the DMRff method(39). We chose this method as it provides decent power for detecting DMRs and without an inflated Type I error rate(40).

We conducted several secondary analyses for the top ten CpG sites from the EWAS in EHBS and ADNI as well as from the meta-analysis. First, we assessed the correlation between the DNAm beta values of the top CpGs across blood and brain tissue using the Blood–Brain Epigenetic Concordance (BECon) tool (46), and the data from Braun et al. (2019) on the Gene Expression Omnibus Database [Accession code GSE111165] (41). To further aid the interpretation of our top associations, we conducted a gene ontology (GO) and KEGG pathway enrichment analysis using the R package missMethyl based on the top 1000 CpG sites with lowest raw p-values for each EWAS and the meta-analyses results (42).

## Results

### Demographics

There were 495 participants from the EHBS and 122 White participants from ADNI (**Table 1**) who met our inclusion criteria of being cognitively normal with available blood DNAm and CSF AD biomarkers. On average, the EHBS participants were about 10 years younger than the ADNI participants (mean age (sd) EHBS White: 62.9 (6.89) years; EHBS Black: 61.7 (8.06) years; ADNI: 74.2 (5.97)) and included more females, particularly among the Black participants (EHBS White: 70.2%; EHBS Black: 82.2%; ADNI: 50.8%). Fewer EHBS participants had a history of tobacco smoking (EHBS White: 30.7%; EHBS Black: 31.1%) compared to ADNI participants (42.6%). EHBS participants had a higher prevalence of the *APOE E4* allele than ADNI participants (**Table 1**).

**Table 1.** Characteristics of the participants from EHBS and ADNI.

|  | EHBS (N=495) |  | ADNI (N=122) |
| --- | --- | --- | --- |
| Race | White (N=450) | Black (N=45) | White (N=122) |
| **A. Characteristics** |  |  |  |
| **Sex** |  |  |  |
| Female | 316 (70.2%) | 37 (82.2%) | 62 (50.8%) |
| Male | 134 (29.8%) | 8 (17.8%) | 60 (49.2%) |
| **Age** |  |  |  |
| Mean (SD) | 62.9 (6.89) | 61.7 (8.06) | 74.2 (5.97) |
| Median [Min, Max] | 63.6 [45.2, 77.0] | 59.5 [50.1, 77.7] | 73.5 [62.0, 89.6] |
| **Smoking** |  |  |  |
| No | 312 (69.3%) | 31 (68.9%) | 70 (57.4%) |
| Yes | 138 (30.7%) | 14 (31.1%) | 52 (42.6%) |
| **APOE4** |  |  |  |
| 0 | 312 (69.3%) | 24 (53.3%) | 94 (77.0%) |
| 1 | 122 (27.1%) | 18 (40.0%) | 25 (20.5%) |
| 2 | 15 (3.3%) | 2 (4.4%) | 3 (2.5%) |
| Missing | 1 (0.2%) | 1 (2.2%) | 0 (0%) |
| **B. CSF AD biomarkers** |  |  |  |
| **tTau** |  |  |  |
| Mean (SD) | 190 (68.0) | 155 (47.5) | 250 (88.1) |
| Median [Min, Max] | 176 [80.0, 555] | 161 [80.2, 282] | 227 [123, 574] |
| **pTau** |  |  |  |
| Mean (SD) | 16.9 (6.72) | 14.4 (4.67) | 22.9 (9.27) |
| Median [Min, Max] | 15.4 [8.00, 61.5] | 14.9 [8.00, 24.5] | 20.0 [10.4, 60.0] |
| **Aβ42/tTau** |  |  |  |
| Mean (SD) | 6.80 (2.04) | 6.71 (1.86) | 5.53 (2.34) |
| Median [Min, Max] | 7.07 [0.692, 11.3] | 6.72 [2.17, 10.5] | 5.71 [0.989, 10.5] |
| **Aβ42+/-** |  |  |  |
| A- | 324 (72.0%) | 23 (51.1%) | 84 (68.9%) |
| A+ | 126 (28.0%) | 22 (48.9%) | 38 (31.1%) |
| **pTau+/-** |  |  |  |
| T- | 365 (81.1%) | 41 (91.1%) | 69 (56.6%) |
| T+ | 85 (18.9%) | 4 (8.9%) | 53 (43.4%) |

In line with their older age, ADNI participants showed more signs of AD-related changes in CSF AD biomarkers in comparison to the EHBS participants. Average levels of tTau and pTau were lower among EHBS participants (EHBS White: mean (sd) tTau: 190 (68.0), pTau: 16.9 (6.72); EHBS Black: mean (sd) tTau: 155 (47.5), pTau: 14.4 (4.67)) than among ADNI participants (ADNI: mean (sd) tTau: 250 (88.1); pTau: 22.9 (9.27)). The distribution of Aβ42+/- was similar across the White EHBS participants and ADNI participants (EHBS White: 28.0% Aβ42+, ADNI: 31.1% Aβ42+) but the proportion of Black EHBS participants who were Aβ42+ was higher than both EHBS White participants and ADNI participants (48.9%).

### Epigenome-wide association study

We investigated whether blood DNAm was associated with CSF Aβ42, pTau, and tTau biomarkers in 617 cognitively normal participants enrolled in EHBS and ADNI. In the EWAS of the White EHBS participants (N=450), no CpG sites were significant at the Bonferroni adjusted p-value<0.05 threshold (equivalent to p< 7.56 x 10^-8^) for any of the AD CSF biomarkers after adjusting for age, sex, smoking history, and estimated cell-type proportions (B lymphocytes, natural killer cells, CD4□+□T lymphocytes, CD8□+□T lymphocytes, monocytes, neutrophils) (**Figure 1**, **Table 2, Figure S1**). At a less stringent EWAS p-value threshold, we found several CpG sites with p-value < 1E-05 for these CSF AD biomarkers and several were found in more than one AD biomarkers with most of the overlap observed between tTau and pTau (**Figure 1F**). For example, cg03586820, which was closest to the gene *SZRD1,* was among the ten most significant CpG sites for tTau (effect estimate: -0.112, p-value: 3.31e-07), pTau (effect estimate: -0.073, p-value: 2.64e-06) and Aβ42/tTau (effect estimate: 0.003, p-value: 3.32e-07). The CpG site cg13422045, assigned to *ARHGEF17,* also showed similar associations across three CSF AD biomarkers (tTau, pTau and pTau+/-), with the strongest associations observed for pTau (effect estimate: -0.010, p-value: 2.76e-06) and pTau+/- (effect estimate: -0.003, p-value: 8.23e-06; Figure 1F). None of the top ten CpG sites with the smallest p-values for the association with the five CSF AD biomarkers could be replicated in ADNI at a nominal p-value threshold of 5%. Similarly, none of the top CpG sites could be replicated in the 45 Black/African American EHBS participants (replication results: **Table S1**)

**Figure 1.**
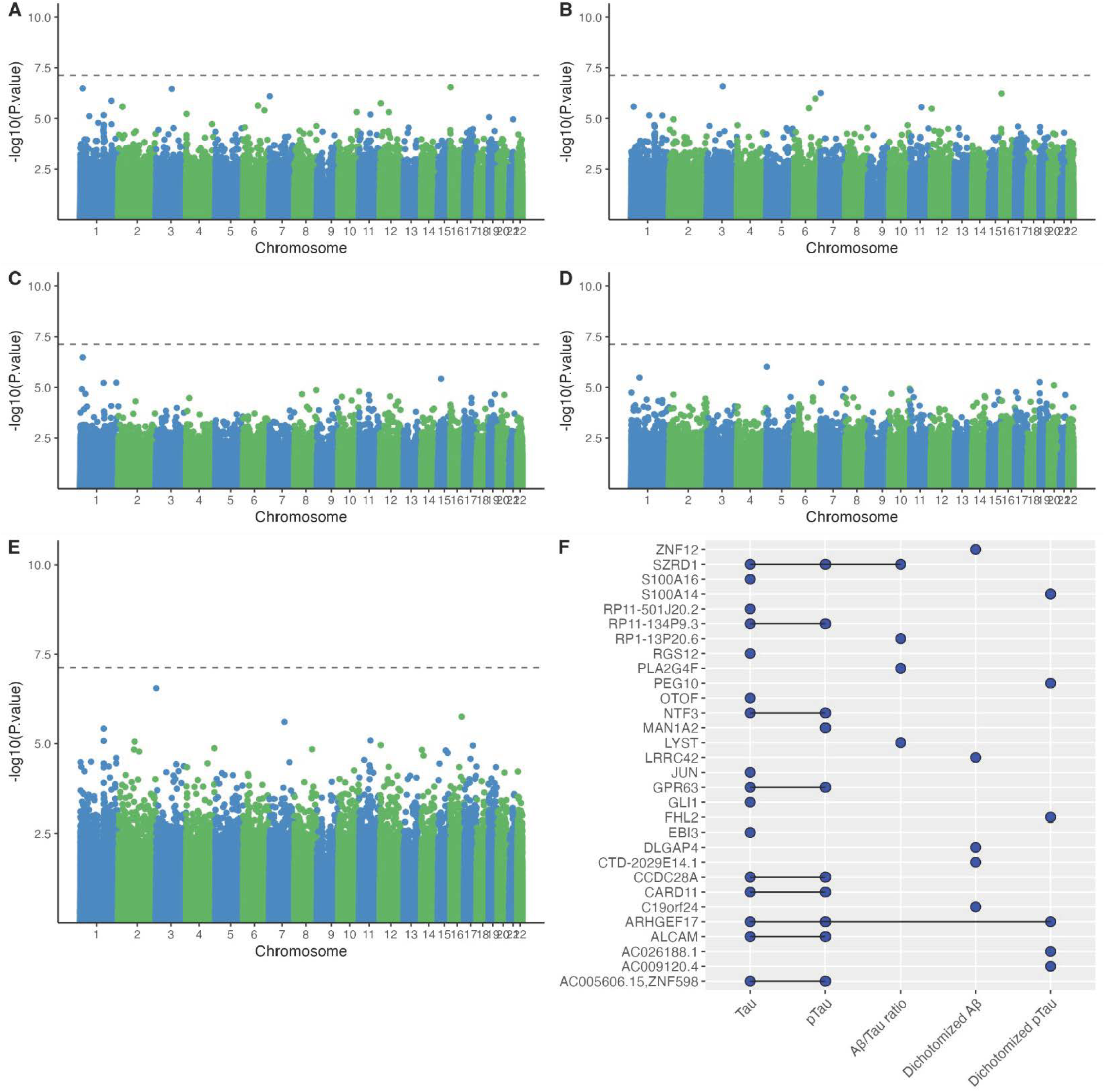
EWAS of AD CSF biomarkers in 450 cognitively normal EHBS participants. Manhattan plots for the association between DNAm beta values and **A.** Tau, **B.** pTau, **C.** Aβ/Tau ratio, **D.** Aβ+/-, **E.** pTau+/-. The dotted line represents the Bonferroni threshold (p = 7.55 x 10^-8^). **F.** UpSet plot showing overlapping associations across the five CSF biomarkers (tTau, pTau, Aβ42/tTau, Aβ42+/-, pTau+/-). A blue dot represents an association between DNAm beta values and the corresponding CSF biomarker with a p-value < 1x 10^-5^for at least one CpG site assigned to the corresponding gene. All associations were adjusted for age at baseline, sex, smoking (with or without smoking history), and estimated cell-type proportions (B lymphocytes, natural killer cells, CD4 + T lymphocytes, CD8 + T lymphocytes, monocytes, neutrophils).

**Table 2.**
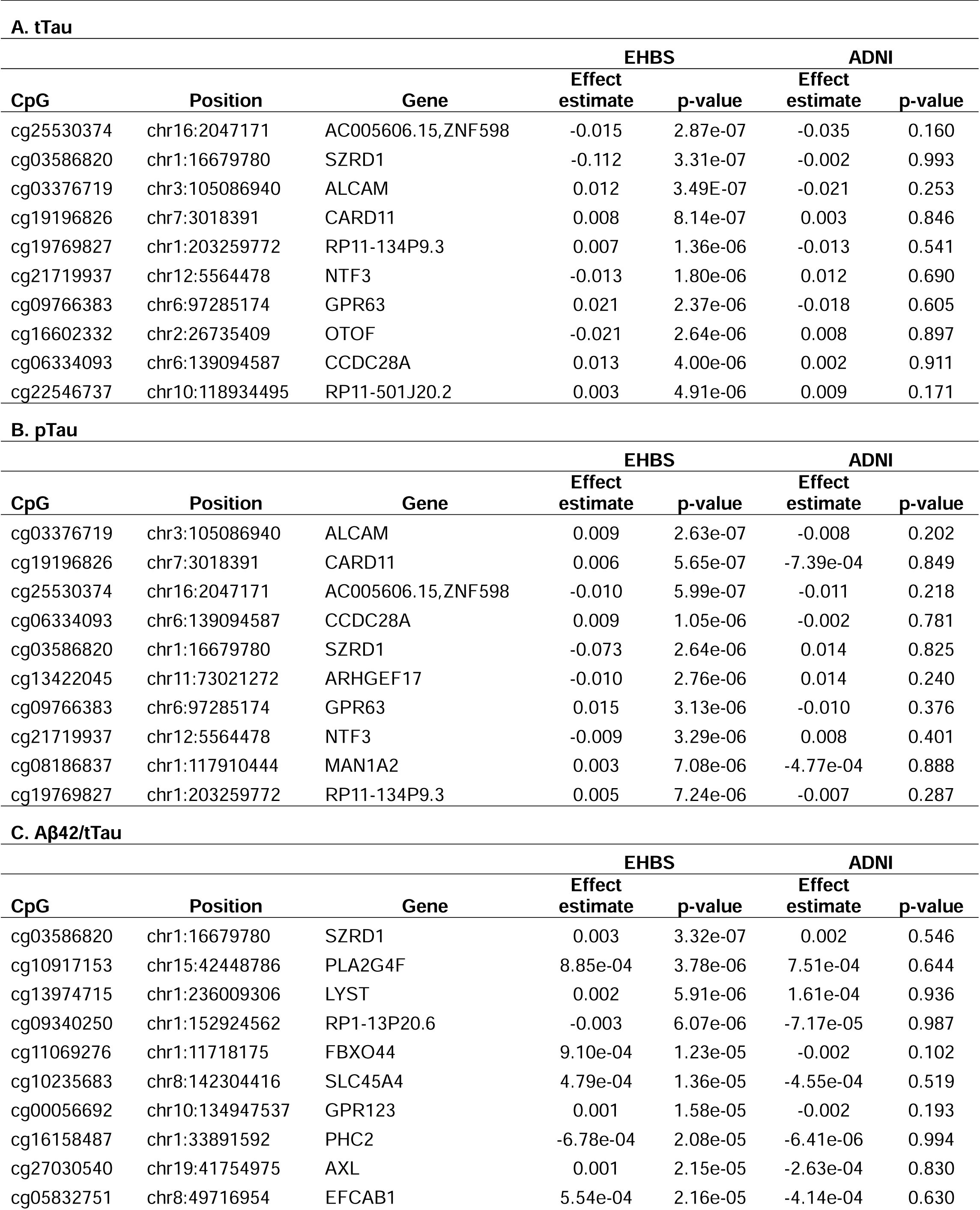
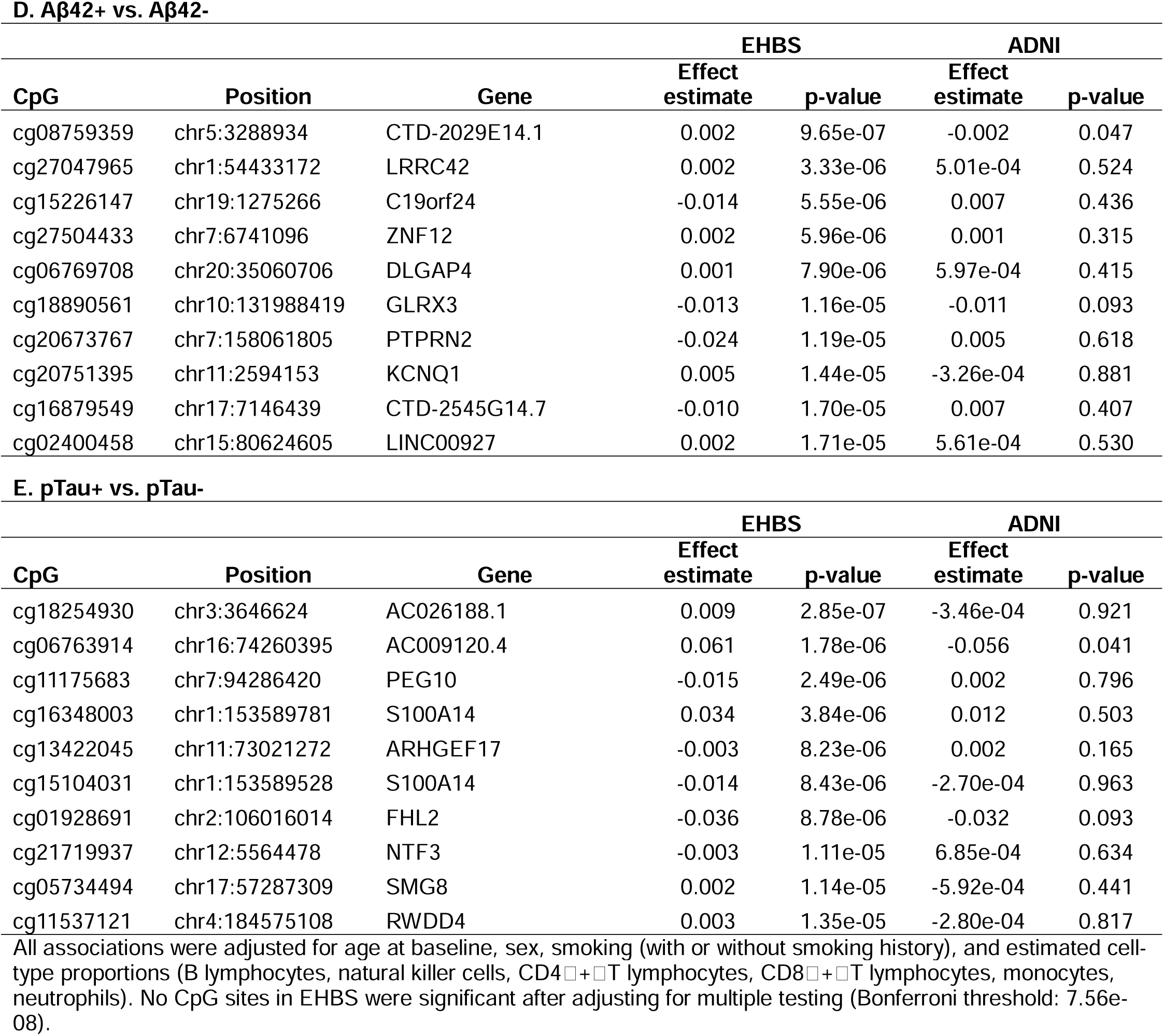
Top 10 CpGs sites from the EWAS of AD CSF biomarkers in EHBS (N=450 White participants) and their replication in ADNI (N=122 White participants).

In ADNI (N=122), two CpG sites were significant at the 5% Bonferroni threshold after adjusting for age, sex, smoking history, and estimated cell-type proportions (B lymphocytes, natural killer cells, CD4□+□T lymphocytes, CD8□+□T lymphocytes, monocytes, neutrophils), including cg21021972 (assigned to the gene *ADAMTS9*) which was significant in Aβ42/tTau (effect estimate: 0.003, p-value: 6.29e-08), and cg17394124 (assigned to the gene *CFH*), which was significant in Aβ42+/- (effect estimate: -0.080, p-value: 7.15e-08) (**Figure 2, Table S2, Figure S2**). However, only one of the top ten CpG sites with the smallest p-values for the association with the five CSF AD biomarkers (and none of the two significant CpG sites) could be replicated in the EHBS at a nominal p-value threshold of 5%. The CpG site cg17394795, which was assigned to the gene *RP11-53B5.1* was among the top ten CpG sites for Aβ42+/- in ADNI (effect estimate: -0.019, p-value: 2.05e-06) and was also nominally significant in the EHBS, although the association was weaker (effect estimate: -0.007, p-value: 0.002).

**Figure 2.**
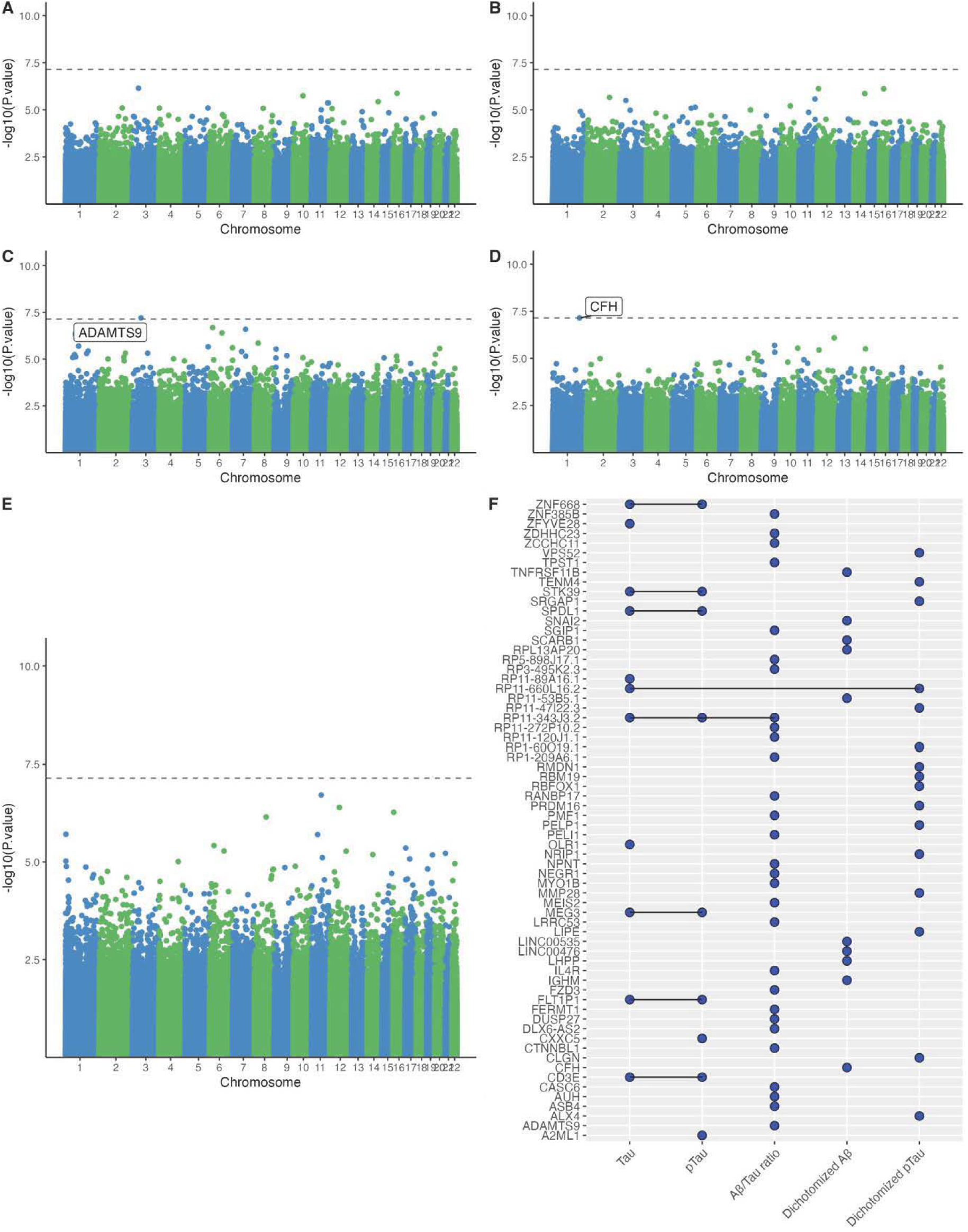
EWAS of AD CSF biomarkers in 122 cognitively normal ADNI participants. Manhattan plots for the association between DNAm beta values and **A.** Tau, **B.** pTau, **C.** Aβ/Tau ratio, **D.** Aβ+/-, **E.** pTau+/-. The dotted line represents the Bonferroni threshold (p = 7.15 x 10^-8^). **F.** UpSet plot showing overlapping associations across the five CSF biomarkers (tTau, pTau, Aβ42/tTau, Aβ42+/-, pTau+/-). A blue dot represents an association between DNAm beta values and the corresponding CSF biomarker with a p-value < 1x 10^-5^for at least one CpG site assigned to the corresponding gene.

We did not find any differentially methylated regions for any of the CSF AD biomarkers in EHBS or ADNI.

### Meta-analysis

In the EWAS meta-analysis of the White EHBS and ADNI participants (N=572), no CpG sites were significant at the Bonferroni adjusted p-value<0.05 threshold (equivalent to p< 7.60 x 10^-8^) for any of the AD CSF biomarkers after adjusting for age, sex, smoking history, and estimated cell-type proportions (B lymphocytes, natural killer cells, CD4 + T lymphocytes, CD8 + T lymphocytes, monocytes, neutrophils) (**Table 3, Figure S3, Figure S4**).

**Table 3.**
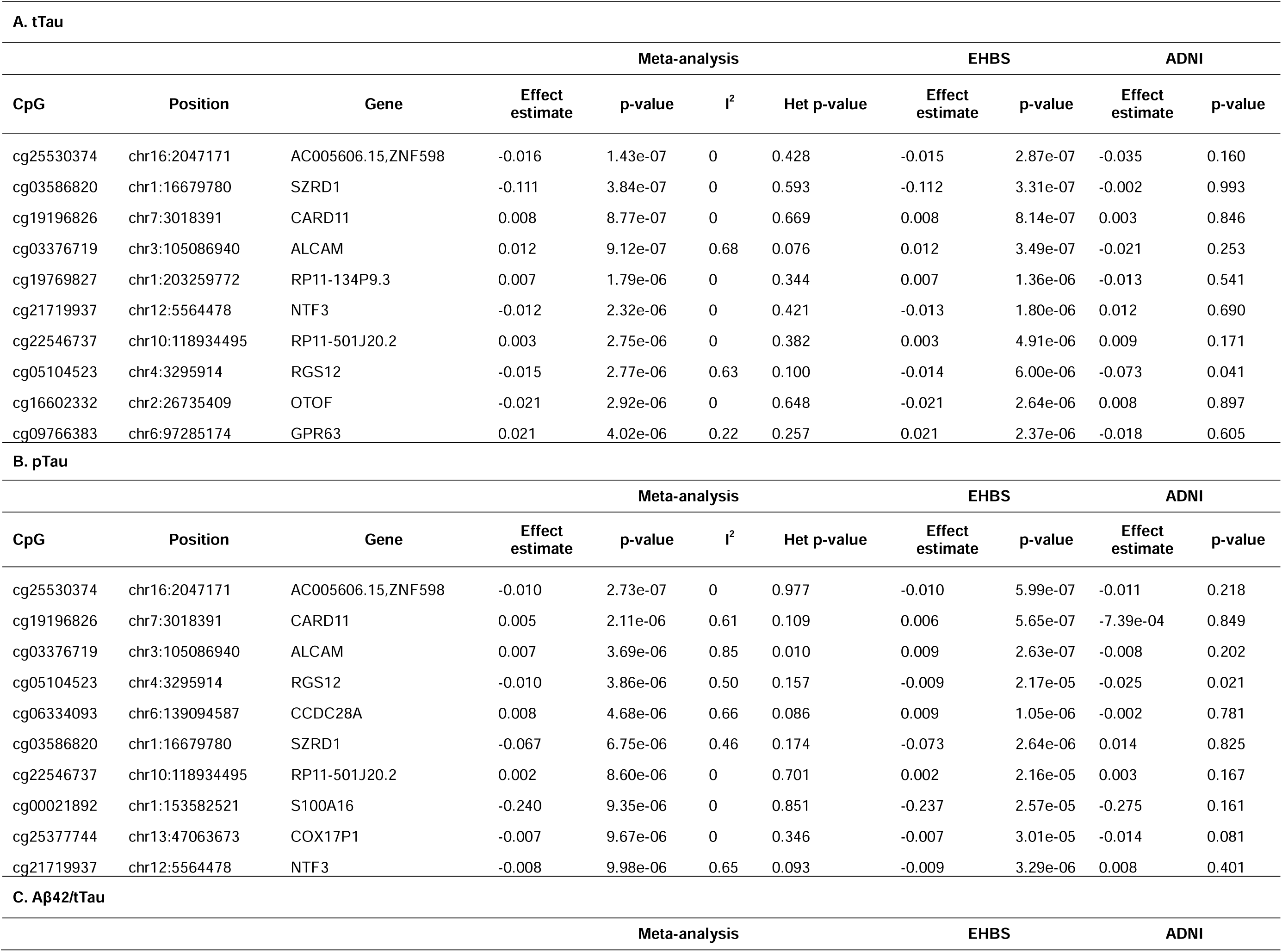
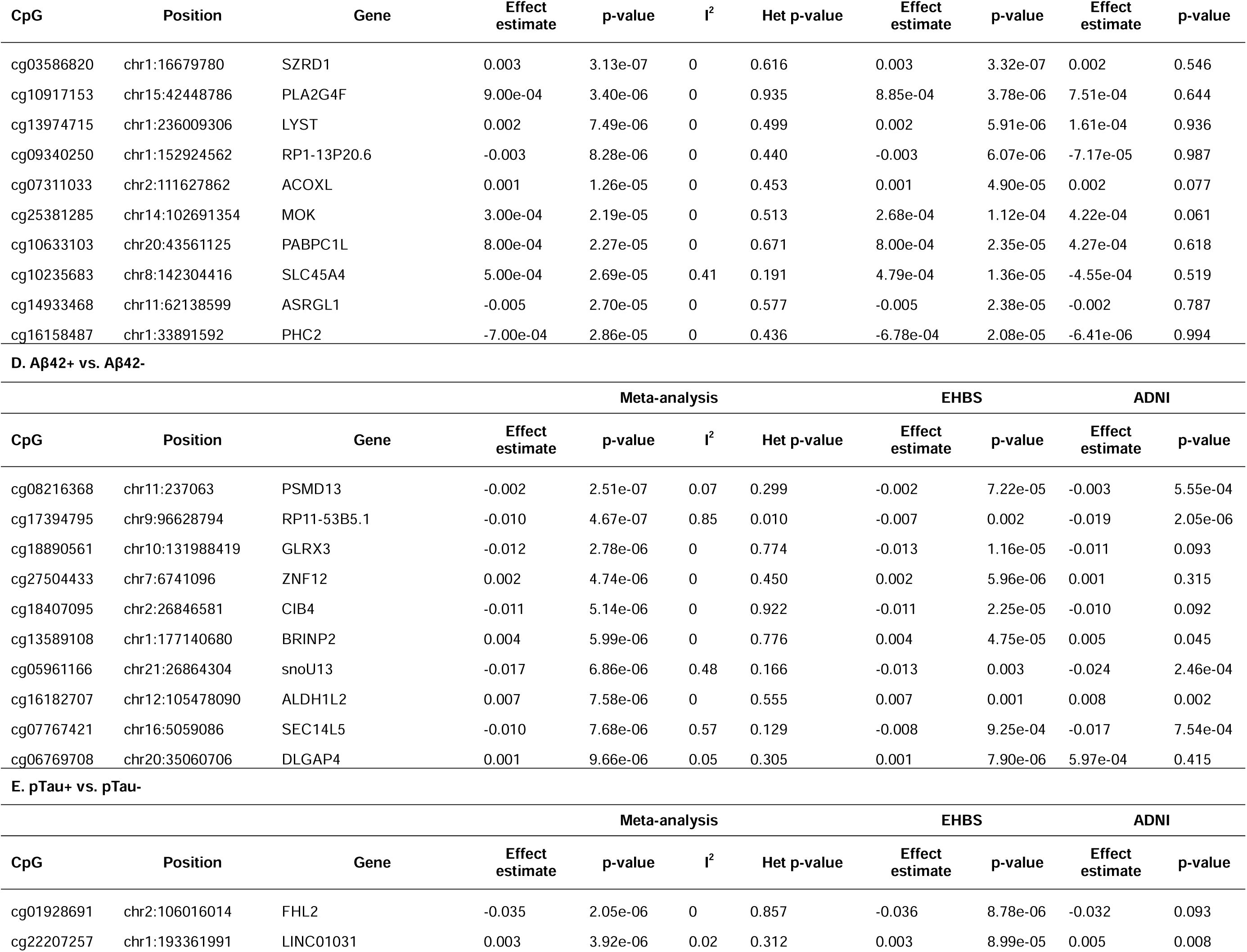
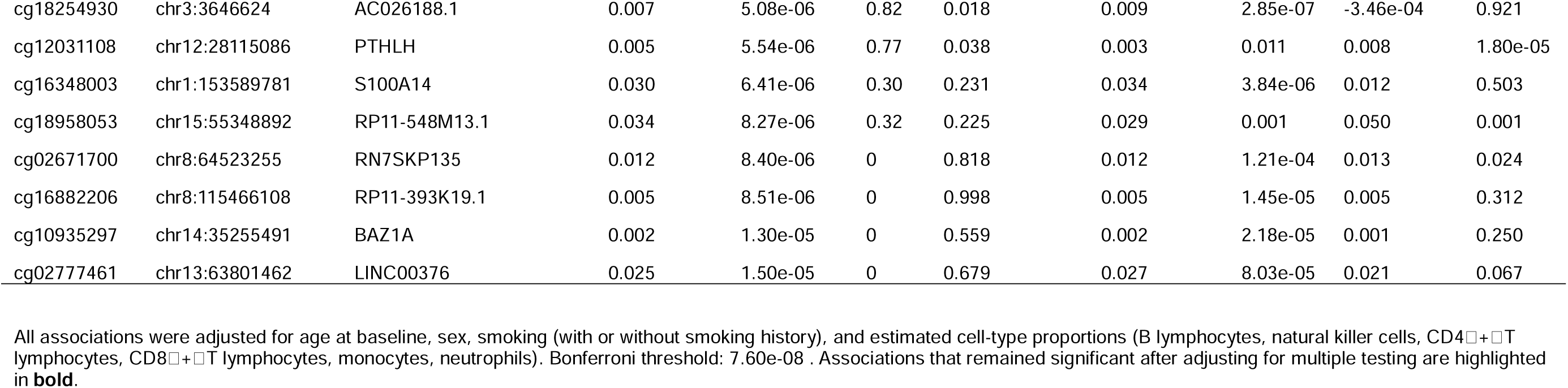
Top 10 CpGs sites from the epigenome-wide meta-analysis of AD CSF biomarkers in 450 cognitively normal individuals from the EHBS and 122 cognitively normal individuals from ADNI.

Since the sample size of the White EHBS participants (N=450) was substantially larger than ADNI White participants (N=122), most of the top associations in the meta-analysis were driven by the EHBS (**Table 3**). For the continuous AD CSF biomarkers, only one of the top 10 CpG sites from the epigenome-wide meta-analysis was at least nominally significant (unadjusted p-value < 0.05) in both cohorts, namely cg05104523 (*RGS12*), which was among the top ten CpG sites for tTau and pTau, For the categorial outcomes of Aβ42+/- and pTau+/-, several of the top 10 CpG sites from the epigenome-wide meta-analysis were at least nominally significant (unadjusted p-value < 0.05) in both cohorts. For Aβ42+/-, six of the top 10 CpG sites were at least nominally significant in both cohorts, including cg08216368 (*PSMD13*), cg17394795 (*RP11-53B5.1*), cg13589108 (*BRINP2*), cg05961166 (*snoU13)*, cg16182707 (*ALDH1L2*) and cg07767421 (*SEC14L5*). For pTau+/-, four of the top 10 CpG sites were at least nominally significant in both cohorts, including cg22207257 (*LINC01031*), cg12031108 (*PTHLH*), cg18958053 (*RP11-548M13.1*) and cg02671700 (*RN7SKP135*).

### Secondary analyses

To further aid the interpretation of our top associations, we performed a gene ontology (GO) and KEGG pathway enrichment analysis based on the top 1000 CpG sites from the EWAS and the meta-analyses with lowest raw p-values. After correction for multiple testing (FDR <0.05), we only identified one KEGG pathway (Parathyroid hormone synthesis, secretion and action) associated with Aβ42/tTau in the EWAS of ADNI. While not statistically significant after correction for multiple testing, the same KEGG pathway was also among the top KEGG pathways for Aβ42/tTau in the EWAS of EHBS (unadjusted p-value = 0.030; 5^th^ smallest p-value) and in the meta-analysis (unadjusted p-value = 0.005; 2^nd^ smallest p-value). We did not identify any GO terms or other KEGG pathways with an overrepresentation of genes containing significantly, differentially methylated CpGs that would indicate an enriched biological pathway. GO terms and KEGG pathways that were nominally significant (raw p<0.05) are included in the supplement (**Tables S3-S11**).

To evaluate the blood–brain concordance for DNAm beta values at our top ten CpG sites, we used the BECon tool and Gene Expression Omnibus Database [Accession code GSE111165]. Several of the top ten CpG sites from the EWAS and the meta-analyses exhibited blood–brain concordance (Tables S3-S5). Among the top 10 CpG sites from the meta-analysis that were at least nominally significant in both cohorts, cg05104523 (*RGS12*, among the top ten CpG sites for tTau and pTau, brain-blood correlation=0.5, p-value=0.022) exhibited blood–brain concordance based on the Gene Expression Omnibus Database.

## Discussion

In the present study, we conducted a blood EWAS of AD CSF biomarkers among 617 cognitively normal participants enrolled in the EHBS and ADNI cohorts. While this is one of the largest EWAS of AD CSF biomarkers and the first that was conducted among cognitively normal individuals, we found little evidence of an association between blood DNAm and AD CSF biomarkers in pre-clinical stages of AD. In the EHBS (N=450 White participants), no CpG sites remained significant at the 5% Bonferroni threshold after adjusting for age, sex, smoking history, and estimated cell-type proportions. In ADNI (N=122 White participants), two CpG sites remained significant at the 5% Bonferroni threshold, but they could not be replicated in the EHBS. Overall, there was little agreement between the top CpG sites from the EWAS of EHBS and of ADNI, respectively, as reflected by little overlap between top CpG sites and no significant findings in the meta-analyses of the two studies. While not statistically significant, a few CpG sites that were either among the top CpG sites for EHBS and ADNI or showed a good agreement across several AD CSF biomarkers are noteworthy and should be further investigated in future studies.

Only two CpG sites were significantly associated with AD CSF biomarkers among 122 cognitively normal participants from the ADNI cohort. Differential DNAm in cg21021972 (assigned to the gene *ADAMTS9*) was significantly associated with Aβ42/tTau, and cg17394124 (assigned to the gene *CFH*) was significantly associated with Aβ42+/-. However, both CpG sites could not be replicated in the EWAS in EHBS, and the meta-analysis of ADNI and EHBS did not identify any significant CpG sites. Our findings could have two potential explanations. First, the absence of a robust association between blood-based DNAm and AD CSF biomarkers in our study might indicate that epigenetic changes in the blood is not a good indicator of pre-clinical stages of AD. This hypothesis is further supported by the weak evidence for an association between blood-based DNAm and cognitive function among cognitively normal individuals. Specifically, a large-scale epigenome-wide meta-analysis of seven measures of cognitive functioning using data from 11 cohorts (N=6809 healthy, older-aged adults) only identified two significant CpG associations with executive function and global cognitive ability (43). Second, even when evaluating epigenetic signatures of AD CSF biomarkers among AD/MCI patients and controls, the most recent EWAS, which included 885 participants from the EMIF-AD study, also did not find strong evidence of an association for CSF amyloid measures and CSF tau variables, as no CpG sites passed the Bonferroni-significance threshold for those measures (22). This study only identified associations between differential DNAm and CSF biomarkers of neuroinflammation (YKL-40) and neurodegeneration (NfL). Another study of 202 ADNI participants identified several FDR-significant loci associated with p-tau181 and Aβ42 (21), but none of those CpG sites could be replicated in the larger EMIF-AD study (22).

While none of the CpG sites in the EHBS EWAS passed the Bonferroni-significance threshold, several CpG sites had p-values < 1e-5 for more than one CSF AD biomarker, with most of the overlap observed between tTau and pTau. For example, cg03586820 (*SZRD1*) was among the ten most significant CpG sites for tTau, pTau and Aβ42/tTau, and cg13422045 (*ARHGEF17*) also showed similar associations across the tau-related CSF AD biomarkers (tTau, pTau and pTau+/-). Interestingly, *ARHGEF17* has been associated with AD Braak stage (44)(45), schizophrenia (46) and mortality (47) in previous studies.

A few CpG sites from our meta-analysis of EHBS and ADNI are noteworthy, as they were among the top 10 CpG sites from the epigenome-wide meta-analysis and at least nominally significant (raw p-value < 0.05) in both cohorts. These include cg05104523 (*RGS12*), which was among the top ten CpG sites for tTau and pTau, cg08216368 (*PSMD13*), cg17394795 (*RP11-53B5.1*), cg13589108 (*BRINP2*), cg05961166 (*snoU13*), cg16182707 (*ALDH1L2*) and cg07767421 (*SEC14L5*), which were among the top CpG sites for Aβ42+/-, and cg22207257 (*LINC01031*), cg12031108 (*PTHLH*), cg18958053 (*RP11-548M13.1*) and cg02671700 (*RN7SKP135*), which were among the top CpG sites for pTau+/-. To our knowledge, *RGS12* is the only gene that has been associated with ADRD previously, in particular with frontotemporal dementia (48). Interestingly, cg05104523 (*RGS12*) also exhibited blood–brain concordance based on the Gene Expression Omnibus Database, suggesting that differential DNAm of cg05104523 in the blood could mirror related changes in the brain.

Our KEGG pathway enrichment analysis identified one KEGG pathway (Parathyroid hormone synthesis, secretion and action) associated with Aβ42/tTau in the EWAS of ADNI, which was also among the top KEGG pathways for Aβ42/tTau in the EWAS of EHBS and in the meta-analysis. Abnormal parathyroid hormone levels play a role in neuronal calcium dysregulation, hypoperfusion and disrupted neuronal signaling and there is some support for a link between parathyroid hormone levels, cognition and dementia (49). A previous systematic review pointed out that mixed findings from previous studies are supported by low and moderate quality data susceptible to confounding effects and limited external validity (49). Given the plausible mechanisms to suggest abnormal parathyroid hormone levels may lead to cognitive dysfunction and an increased risk of dementia, our study is an important contribution for disentangling this relationship.

Our study has several strengths. Most importantly, the EHBS is one of the largest prospective cohort studies with CSF samples from cognitively normal individuals. The level of depth in our outcome assessment, underscored by the inclusion of a substantial sample size with CSF measurements, a highly invasive and challenging-to-obtain biological fluid, provides a rare and valuable opportunity to understand potential associations between differential DNAm and AD CSF biomarkers among cognitively normal individuals. Furthermore, our study is the first study of DNAm and AD CSF biomarkers that attempted a replication of findings in an independent cohort, namely ADNI, thus strengthening the robustness of our conclusions.

In addition to its strengths, our study has several limitations. First, the temporal sequence between differential DNAm and AD CSF biomarkers could not be clearly defined because both were assessed in blood samples collected at the same study visit. Furthermore, while all our study participants were cognitively normal, i.e., without a diagnosis of cognitive impairment at blood draw, there were substantial differences between EHBS and ADNI participants related to the study design and the demographics. EHBS participants were about 10 years younger than the ADNI participants and overall healthier. In addition, there were only 122 cognitively normal ADNI participants with DNAm data (vs. 450 EHBS participants), which might have contributed to the lack of replication across the two cohorts. Another limitation of our study is the use of whole blood for DNAm profiling. Although we have tried to account for this by including cell proportions as covariates in our analyses, future research using DNA isolated from specific cell types would enable the identification of cell type-specific signatures related to the AD CSF biomarkers. Another limitation of our study is that we had to restrict our main analyses to White participants and none of our findings could be replicated in our smaller population of Black/African American EHBS participants of the EHBS.

In conclusion, our EWAS of blood-based DNAm and AD CSF biomarkers among 617 cognitively normal participants enrolled in the EHBS and the ADNI cohort showed only weak evidence of an association between differential DNAm and AD CSF biomarkers assessed to evaluate pre-clinical stages of AD. Future studies should include additional biomarkers, e.g., CSF biomarkers of neuroinflammation (YKL-40) and neurodegeneration (NfL) or blood-based AD biomarkers, which might show a stronger correlation between blood-based DNAm.

## Data Availability

All data produced in the present study are available upon reasonable request to the authors

## Acknowledgments

We gratefully acknowledge the research volunteers and staff of the Goizueta Alzheimer’s Disease Research Center at Emory University, and Emory Healthy Brain Study for their participation and contributions. This work was supported by I01 BX003853 (APW), I01 BX005686 (APW), IK4 BX005219 (APW), P30 AG066511 (AIL, JJL), R01 AG056533 (APW, TSW), R01 AG070937 (JJL), R01 AG072120 (APW, TSW), R01 AG075827 (APW, TSW), R01 AG079170 (AH, TSW), U01 AG046161 (AIL), U01 AG061356 (AIL), U01 AG061357 (AIL), U01 AG088425 (AH, TSW), R01AG087250 (AH)

## Consent Statement

The Emory Healthy Brain Study was approved by the Institutional Review Board of Emory University Medical Center. All participants provided written informed consent.

