## Supplementary Tables and Figures for "Epigenome-wide association study of cerebrospinal fluid-based biomarkers of Alzheimer’s disease in cognitively normal individuals"

| **Table S1. Top 10 CpGs sites from the epigenome-wide association study of cerebrospinal fluid-based (CSF) biomarkers of Alzheimer’s disease in the White participants of the Emory Healthy Brain Study (EHBS; main analysis, N=450) and their replication in the Black/African American (AA) participants of the EHBS (N=45).** | | | | | | |
| --- | --- | --- | --- | --- | --- | --- |
| **A. tTau** | | | | | | |
|  |  |  | **EHBS White** | | **EHBS Black/AA** | |
| **CpG** | **Position** | **Gene** | **Effect estimate** | **p-value** | **Effect estimate** | **p-value** |
| cg25530374 | chr16:2047171 | AC005606.15,ZNF598 | -0.015 | 2.87e-07 | 0.00145 | 0.003 |
| cg03586820 | chr1:16679780 | SZRD1 | -0.112 | 3.31e-07 | -0.00599 | 0.349 |
| cg03376719 | chr3:105086940 | ALCAM | 0.012 | 3.49E-07 | -0.00057 | 0.320 |
| cg19196826 | chr7:3018391 | CARD11 | 0.008 | 8.14e-07 | -0.00008 | 0.863 |
| cg19769827 | chr1:203259772 | RP11-134P9.3 | 0.007 | 1.36e-06 | 0.00028 | 0.591 |
| cg21719937 | chr12:5564478 | NTF3 | -0.013 | 1.80e-06 | 0.00105 | 0.181 |
| cg09766383 | chr6:97285174 | GPR63 | 0.021 | 2.37e-06 | -0.00015 | 0.833 |
| cg16602332 | chr2:26735409 | OTOF | -0.021 | 2.64e-06 | 0.00111 | 0.214 |
| cg06334093 | chr6:139094587 | CCDC28A | 0.013 | 4.00e-06 | 0.00005 | 0.956 |
| cg22546737 | chr10:118934495 | RP11-501J20.2 | 0.003 | 4.91e-06 | -0.00038 | 0.084 |
| **B. pTau** | | | | | | |
|  |  |  | **EHBS White** | | **EHBS Black/AA** | |
| **CpG** | **Position** | **Gene** | **Effect estimate** | **p-value** | **Effect estimate** | **p-value** |
| cg03376719 | chr3:105086940 | ALCAM | 0.009 | 2.63e-07 | -0.00128 | 0.519 |
| cg19196826 | chr7:3018391 | CARD11 | 0.006 | 5.65e-07 | -0.00089 | 0.572 |
| cg25530374 | chr16:2047171 | AC005606.15,ZNF598 | -0.010 | 5.99e-07 | 0.00462 | 0.005 |
| cg06334093 | chr6:139094587 | CCDC28A | 0.009 | 1.05e-06 | -0.00054 | 0.867 |
| cg03586820 | chr1:16679780 | SZRD1 | -0.073 | 2.64e-06 | -0.03050 | 0.239 |
| cg13422045 | chr11:73021272 | ARHGEF17 | -0.010 | 2.76e-06 | 0.00302 | 0.221 |
| cg09766383 | chr6:97285174 | GPR63 | 0.015 | 3.13e-06 | -0.00103 | 0.650 |
| cg21719937 | chr12:5564478 | NTF3 | -0.009 | 3.29e-06 | 0.00321 | 0.240 |
| cg08186837 | chr1:117910444 | MAN1A2 | 0.003 | 7.08e-06 | -0.00053 | 0.746 |
| cg19769827 | chr1:203259772 | RP11-134P9.3 | 0.005 | 7.24e-06 | 0.00027 | 0.882 |
| **C. Aβ42/tTau** | | | | | | |
|  |  |  | **EHBS White** | | **EHBS Black/AA** | |
| **CpG** | **Position** | **Gene** | **Effect estimate** | **p-value** | **Effect estimate** | **p-value** |
| cg03586820 | chr1:16679780 | SZRD1 | 0.003 | 3.32e-07 | -0.00015 | 0.924 |
| cg10917153 | chr15:42448786 | PLA2G4F | 8.85e-04 | 3.78e-06 | -0.00053 | 0.485 |
| cg13974715 | chr1:236009306 | LYST | 0.002 | 5.91e-06 | 0.00197 | 0.046 |
| cg09340250 | chr1:152924562 | RP1-13P20.6 | -0.003 | 6.07e-06 | 0.00043 | 0.818 |
| cg11069276 | chr1:11718175 | FBXO44 | 9.10e-04 | 1.23e-05 | -0.00084 | 0.050 |
| cg10235683 | chr8:142304416 | SLC45A4 | 4.79e-04 | 1.36e-05 | -0.00039 | 0.110 |
| cg00056692 | chr10:134947537 | GPR123 | 0.001 | 1.58e-05 | -0.00022 | 0.799 |
| cg16158487 | chr1:33891592 | PHC2 | -6.78e-04 | 2.08e-05 | 0.00114 | 0.049 |
| cg27030540 | chr19:41754975 | AXL | 0.001 | 2.15e-05 | 0.00037 | 0.605 |
| cg05832751 | chr8:49716954 | EFCAB1 | 5.54e-04 | 2.16e-05 | 0.00028 | 0.634 |
| **D. Aβ42+/-** | | | | | | |
|  |  |  | **EHBS White** | | **EHBS Black/AA** | |
| **CpG** | **Position** | **Gene** | **Effect estimate** | **p-value** | **Effect estimate** | **p-value** |
| cg08759359 | chr5:3288934 | CTD-2029E14.1 | 0.002 | 9.65e-07 | -0.00164 | 0.251 |
| cg27047965 | chr1:54433172 | LRRC42 | 0.002 | 3.33e-06 | -0.00006 | 0.965 |
| cg15226147 | chr19:1275266 | C19orf24 | -0.014 | 5.55e-06 | 0.01986 | 0.036 |
| cg27504433 | chr7:6741096 | ZNF12 | 0.002 | 5.96e-06 | 0.00015 | 0.920 |
| cg06769708 | chr20:35060706 | DLGAP4 | 0.001 | 7.90e-06 | 0.00065 | 0.614 |
| cg18890561 | chr10:131988419 | GLRX3 | -0.013 | 1.16e-05 | 0.01893 | 0.111 |
| cg20673767 | chr7:158061805 | PTPRN2 | -0.024 | 1.19e-05 | 0.00244 | 0.890 |
| cg20751395 | chr11:2594153 | KCNQ1 | 0.005 | 1.44e-05 | 0.00262 | 0.573 |
| cg16879549 | chr17:7146439 | CTD-2545G14.7 | -0.010 | 1.70e-05 | -0.01413 | 0.085 |
| cg02400458 | chr15:80624605 | LINC00927 | 0.002 | 1.71e-05 | 0.00065 | 0.732 |
| **E. pTau+/-** | | | | | | |
|  |  |  | **EHBS White** | | **EHBS Black/AA** | |
| **CpG** | **Position** | **Gene** | **Effect estimate** | **p-value** | **Effect estimate** | **p-value** |
| cg18254930 | chr3:3646624 | AC026188.1 | 0.009 | 2.85e-07 | 0.01922 | 0.374 |
| cg06763914 | chr16:74260395 | AC009120.4 | 0.061 | 1.78e-06 | 0.00819 | 0.913 |
| cg11175683 | chr7:94286420 | PEG10 | -0.015 | 2.49e-06 | -0.00599 | 0.770 |
| cg16348003 | chr1:153589781 | S100A14 | 0.034 | 3.84e-06 | 0.01669 | 0.556 |
| cg13422045 | chr11:73021272 | ARHGEF17 | -0.003 | 8.23e-06 | 0.00032 | 0.939 |
| cg15104031 | chr1:153589528 | S100A14 | -0.014 | 8.43e-06 | 0.00043 | 0.972 |
| cg01928691 | chr2:106016014 | FHL2 | -0.036 | 8.78e-06 | 0.05383 | 0.141 |
| cg21719937 | chr12:5564478 | NTF3 | -0.003 | 1.11e-05 | 0.00603 | 0.039 |
| cg05734494 | chr17:57287309 | SMG8 | 0.002 | 1.14e-05 | -0.00508 | 0.007 |
| cg11537121 | chr4:184575108 | RWDD4 | 0.003 | 1.35e-05 | 0.00546 | 0.093 |
| All associations were adjusted for age at baseline, sex, smoking (with or without smoking history), and estimated cell-type proportions (B lymphocytes, natural killer cells, CD4 + T lymphocytes, CD8 + T lymphocytes, monocytes, neutrophils). No CpG sites in EHBS remained significant after adjusting for multiple testing (Bonferroni threshold: 7.56e-08). | | | | | | |

| **Table S2. Top 10 CpGs sites from the epigenome-wide association study of cerebrospinal fluid-based (CSF) biomarkers of Alzheimer’s disease in ADNI (N=122) and their replication in the Emory Healthy Brain Study (EHBS, N=450 White participants).** | | | | | | |
| --- | --- | --- | --- | --- | --- | --- |
| **A. tTau** | | | | | | |
|  |  |  | **ADNI** | | **EHBS** | |
| **CpG** | **Position** | **Gene** | **Effect estimate** | **p-value** | **Effect estimate** | **p-value** |
| cg08522625 | chr3:46135532 | FLT1P1 | -0.282 | 7.10e-07 | -0.004 | 0.510 |
| cg10071605 | chr16:31072718 | ZNF668 | -0.202 | 1.33e-06 | -0.005 | 0.458 |
| cg17162734 | chr10:71337586 | RP11-343J3.2 | 0.104 | 1.80e-06 | 0.002 | 0.568 |
| cg13468451 | chr14:101315775 | MEG3 | -0.165 | 3.67e-06 | 0.005 | 0.252 |
| cg12833422 | chr11:118166049 | CD3E | 0.930 | 4.28e-06 | -0.014 | 0.668 |
| cg19026692 | chr2:168947421 | STK39 | 0.061 | 7.98e-06 | 0.002 | 0.320 |
| cg18581037 | chr5:169010289 | SPDL1 | 0.097 | 8.00e-06 | 0.002 | 0.494 |
| cg10604715 | chr4:2279733 | ZFYVE28 | -0.217 | 8.16e-06 | -0.009 | 0.162 |
| cg22868272 | chr8:65936215 | RP11-89A16.1 | -0.113 | 8.34e-06 | 0.001 | 0.764 |
| cg19518069 | chr12:10320770 | OLR1 | -0.900 | 8.47e-06 | -0.024 | 0.230 |
| **B. pTau** | | | | | | |
|  |  |  | **ADNI** | | **EHBS** | |
| **CpG** | **Position** | **Gene** | **Effect estimate** | **p-value** | **Effect estimate** | **p-value** |
| cg21416544 | chr12:8986988 | A2ML1 | -0.226 | 7.51e-07 | 0.013 | 0.394 |
| cg10071605 | chr16:31072718 | ZNF668 | -0.069 | 7.67e-07 | -0.003 | 0.484 |
| cg13468451 | chr14:101315775 | MEG3 | -0.053 | 1.38e-06 | 0.004 | 0.165 |
| cg19026692 | chr2:168947421 | STK39 | 0.021 | 2.19e-06 | 0.001 | 0.254 |
| cg12833422 | chr11:118166049 | CD3E | 0.317 | 2.66e-06 | -0.007 | 0.734 |
| cg08522625 | chr3:46135532 | FLT1P1 | -0.087 | 3.18e-06 | -0.003 | 0.469 |
| cg17162734 | chr10:71337586 | RP11-343J3.2 | 0.032 | 6.20e-06 | 0.001 | 0.594 |
| cg18581037 | chr5:169010289 | SPDL1 | 0.030 | 7.33e-06 | 0.001 | 0.516 |
| cg11910223 | chr5:139060821 | CXXC5 | -0.069 | 8.22e-06 | 0.010 | 0.019 |
| cg22868272 | chr8:65936215 | RP11-89A16.1 | -0.036 | 1.01e-05 | 4.33e-04 | 0.885 |
| **C.** **Aβ42/tTau** | | | | | | |
|  |  |  | **ADNI** | | **EHBS** | |
| **CpG** | **Position** | **Gene** | **Effect estimate** | **p-value** | **Effect estimate** | **p-value** |
| cg21021972 | chr3:64613818 | ADAMTS9 | **0.003** | **6.29e-08** | -2.27e-04 | 0.300 |
| cg04434425 | chr6:22929545 | RP1-209A6.1 | -0.002 | 2.06e-07 | 2.08e-05 | 0.879 |
| cg09034302 | chr7:95108238 | ASB4 | 0.020 | 2.55e-07 | -1.96e-04 | 0.860 |
| cg01028017 | chr6:92363854 | CASC6 | 0.018 | 4.02e-07 | 0.001 | 0.125 |
| cg04985396 | chr1:72749735 | NEGR1 | -0.003 | 4.60e-07 | 2.08e-04 | 0.109 |
| cg16729690 | chr8:28351217 | FZD3 | -0.001 | 1.39e-06 | 5.03e-05 | 0.317 |
| cg03455845 | chr1:96840865 | RP5-898J17.1 | 0.005 | 2.00e-06 | -4.08e-04 | 0.166 |
| cg27234746 | chr5:170579510 | RANBP17 | 0.005 | 2.20e-06 | -1.22e-04 | 0.649 |
| cg24738424 | chr6:169364882 | RP3-495K2.3 | 0.006 | 2.47e-06 | 2.12e-04 | 0.606 |
| cg10037002 | chr20:36378118 | CTNNBL1 | 0.011 | 2.71e-06 | -0.001 | 0.065 |
| **D.** **Aβ42+/-** | | | | | | |
|  |  |  | **ADNI** | | **EHBS** | |
| **CpG** | **Position** | **Gene** | **Effect estimate** | **p-value** | **Effect estimate** | **p-value** |
| cg17394124 | chr1:196712668 | CFH | **-0.080** | **7.15e-08** | -0.008 | 0.310 |
| cg08770004 | chr12:125170382 | SCARB1 | 0.023 | 8.13e-07 | 0.005 | 0.045 |
| cg17394795 | chr9:96628794 | RP11-53B5.1 | -0.019 | 2.05e-06 | -0.007 | 0.002 |
| cg04903883 | chr14:106321551 | IGHM | -0.012 | 3.12e-06 | -6.26e-04 | 0.662 |
| cg06014227 | chr12:13028038 | RPL13AP20 | -0.009 | 3.61e-06 | 7.30e-04 | 0.470 |
| cg14123436 | chr9:98544579 | LINC00476 | -0.005 | 4.70e-06 | 1.18e-04 | 0.853 |
| cg06869529 | chr8:94594936 | LINC00535 | -0.014 | 5.18e-06 | 9.66e-04 | 0.673 |
| cg16624991 | chr8:119931917 | TNFRSF11B | -0.039 | 6.95e-06 | 3.01e-04 | 0.958 |
| cg12252090 | chr8:49833279 | SNAI2 | 0.056 | 9.97e-06 | 0.002 | 0.755 |
| **E. pTau+/-** | | | | | | |
|  |  |  | **ADNI** | | **EHBS** | |
| **CpG** | **Position** | **Gene** | **Effect estimate** | **p-value** | **Effect estimate** | **p-value** |
| cg07487535 | chr11:71159905 | RP11-660L16.2 | -0.005 | 1.94e-07 | 4.16e-05 | 0.950 |
| cg16666973 | chr12:64492812 | SRGAP1 | 0.003 | 4.01e-07 | -0.001 | 0.102 |
| cg06233376 | chr16:6532764 | RBFOX1 | 0.109 | 5.35e-07 | -0.011 | 0.351 |
| cg15253520 | chr8:87521369 | RMDN1 | -0.055 | 7.06e-07 | 0.008 | 0.252 |
| cg17104202 | chr1:3162404 | PRDM16 | 0.005 | 1.95e-06 | -0.001 | 0.145 |
| cg05853285 | chr11:44339621 | ALX4 | 0.028 | 1.99e-06 | -1.96e-04 | 0.962 |
| cg00879843 | chr6:33239764 | VPS52 | -0.002 | 3.80e-06 | 5.18e-04 | 0.187 |
| cg14389924 | chr17:4606729 | PELP1 | 0.031 | 4.42e-06 | 0.003 | 0.424 |
| cg00611605 | chr6:107236467 | RP1-60O19.1 | 0.080 | 5.24e-06 | 0.005 | 0.628 |
| cg03459510 | chr12:114300370 | RBM19 | -0.028 | 5.28e-06 | -2.54e-05 | 0.996 |
| All associations were adjusted for age at baseline, sex, smoking (with or without smoking history), and estimated cell-type proportions (B lymphocytes, natural killer cells, CD4 + T lymphocytes, CD8 + T lymphocytes, monocytes, neutrophils). Bonferroni threshold in ADNI: 7.15e-08. Associations that remained significant after adjusting for multiple testing are highlighted in **bold**. | | | | | | |

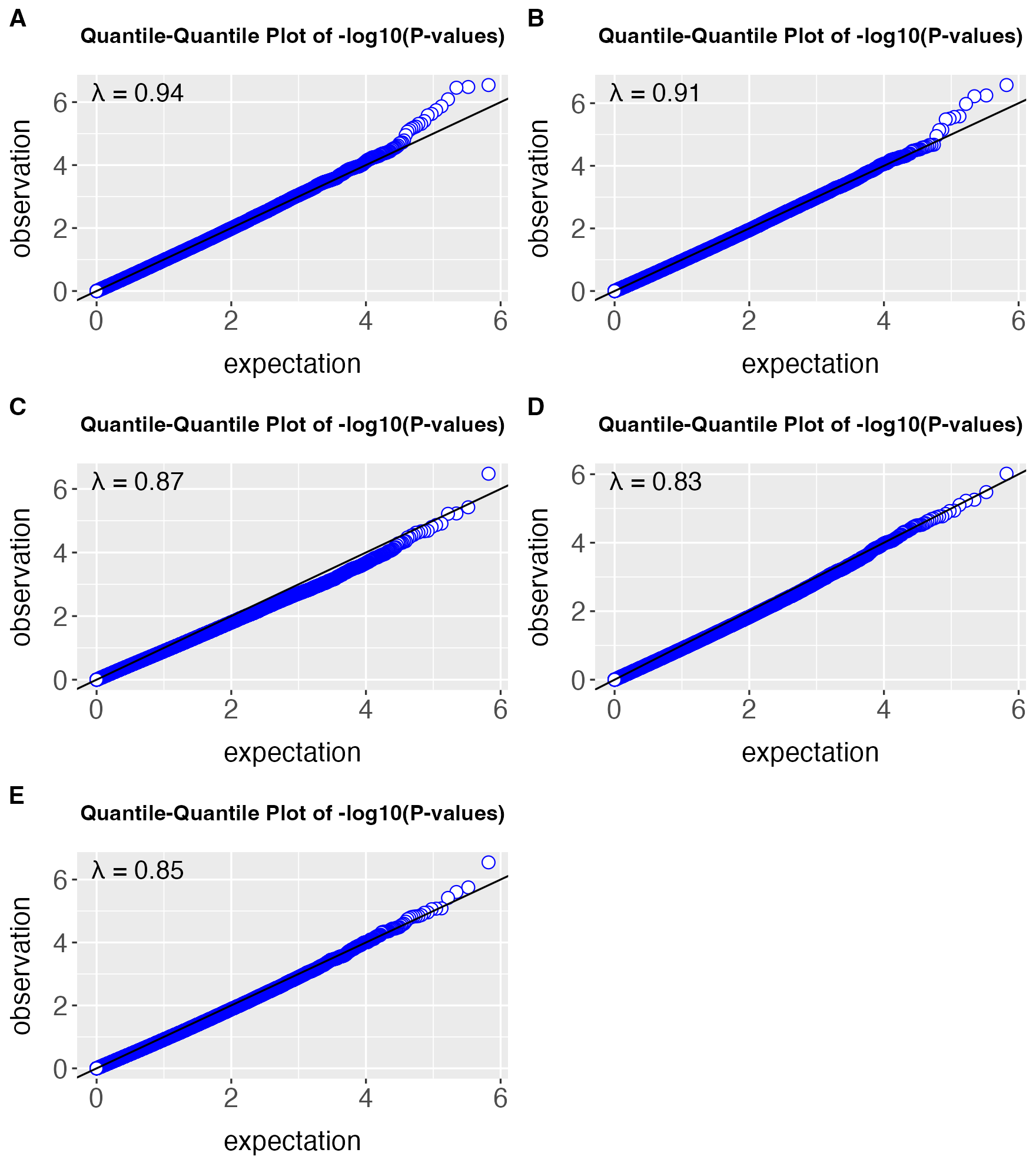

**Figure S1. A.** QQ-plot of EWAS on Tau in EHBS. **B.** QQ-plot of EWAS on pTau in EHBS. **C.** QQ-plot of EWAS on ratio of A-beta and Tau in EHBS. **D.** QQ-plot of EWAS on dichotomized A-beta in EHBS. **E.** QQ-plot of EWAS on dichotomized pTau in EHBS.

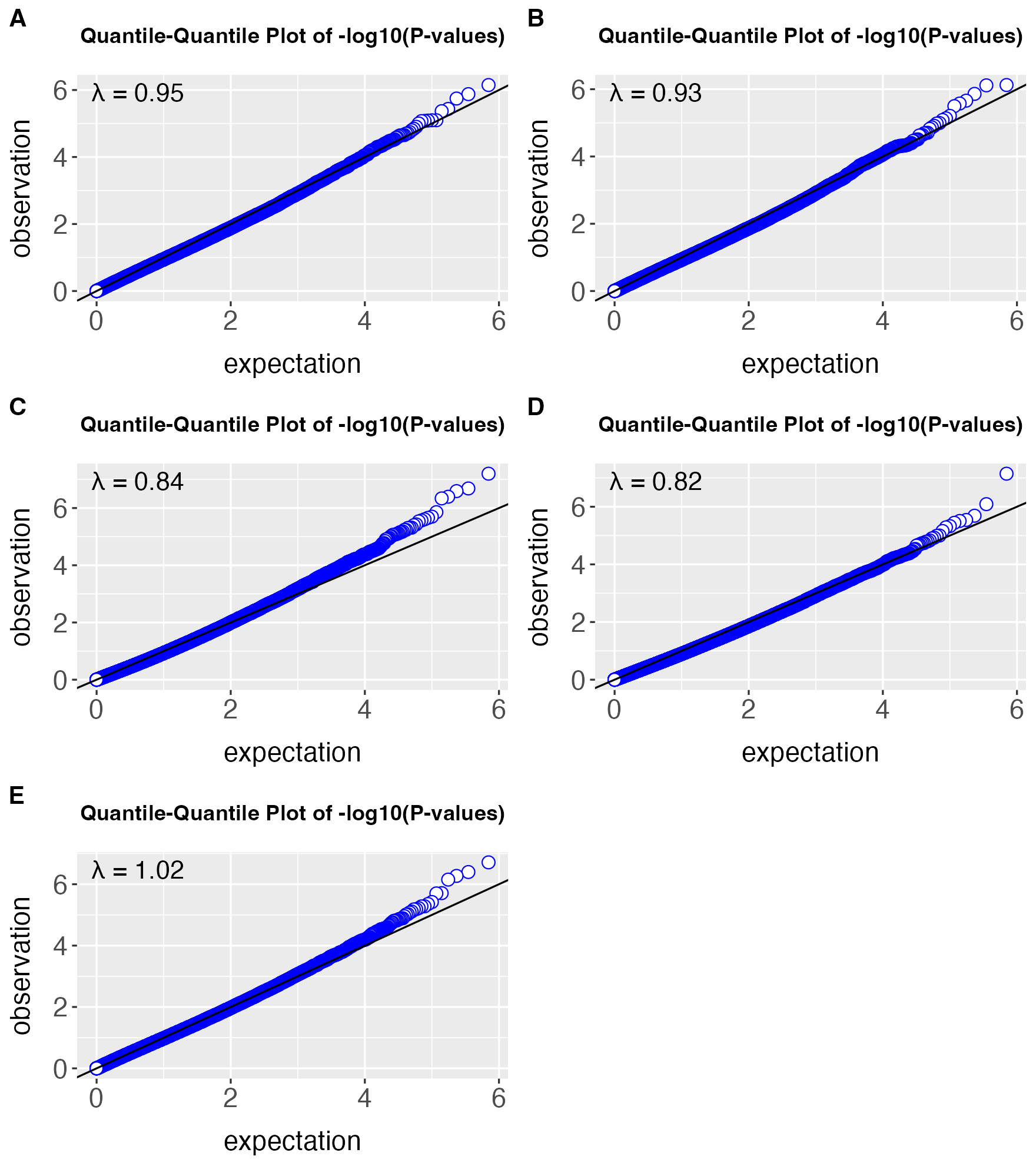

**Figure S2. A.** QQ-plot of EWAS on Tau in ADNI. **B.** QQ-plot of EWAS on pTau in ADNI. **C.** QQ-plot of EWAS on ratio of A-beta and Tau in ADNI. **D.** QQ-plot of EWAS on dichotomized A-beta in ADNI. **E.** QQ-plot of EWAS on dichotomized pTau in ADNI.

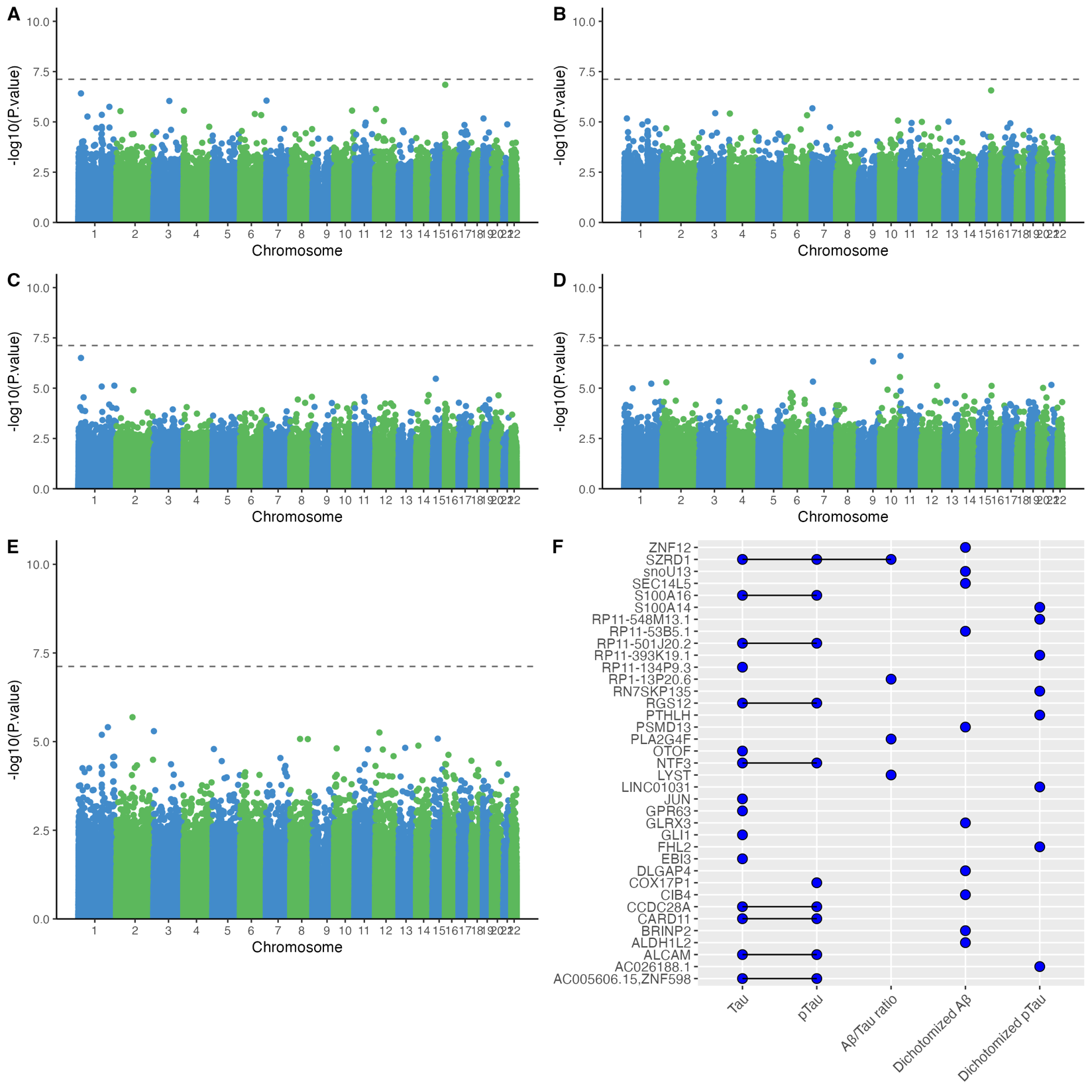
**Figure S3**. **Meta-analysis of CSF biomarkers EWAS of cognitively normal individuals from EHBS and ADNI.** Manhattan plots for the association between DNAm beta values and **A.** tTau, **B.** pTau, **C.** Aβ42/tTau, **D.** Aβ42+/-, **E.** pTau+/-. The dotted line represents the Bonferroni threshold ($p=7.60\times{10}^{-8}$). **F.** UpSet plot showing overlapping associations across the five CSF biomarkers (tTau, pTau, Aβ42/tTau, Aβ42+/-, pTau+/-). A blue dot represents an association between DNAm beta values and the corresponding CSF biomarker with a p-value < $1\times{10}^{-5}$ for at least one CpG site assigned to the corresponding gene. All associations were adjusted for age at baseline, sex, smoking (with or without smoking history), and estimated cell-type proportions (B lymphocytes, natural killer cells, CD4 + T lymphocytes, CD8 + T lymphocytes, monocytes, neutrophils).

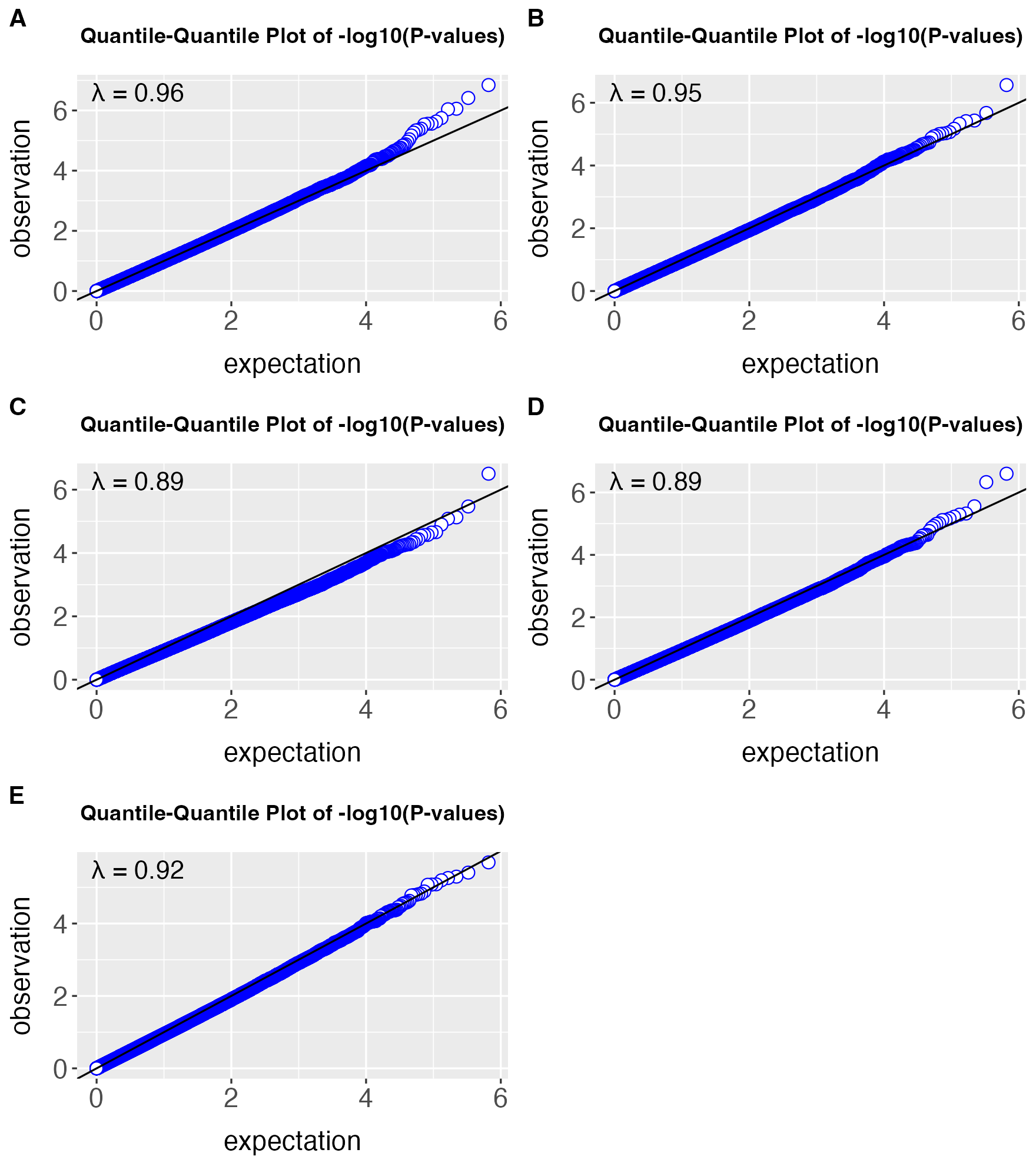

**Figure S4.**  **Epigenome-wide meta-analysis of cerebrospinal fluid-based (CSF) biomarkers of Alzheimer’s disease in in 450 cognitively normal individuals from the Emory Healthy Brain Study (EHBS) and 122 cognitively normal individuals from ADNI. A.** QQ-plot of meta-analysis on Tau. **B.** QQ-plot of meta-analysis on pTau. **C.** QQ-plot of meta-analysis on ratio of A-beta and Tau. **D.** QQ-plot of meta-analysis on dichotomized A-beta. **E.** QQ-plot of meta-analysis on dichotomized pTau.

| **Table S3. Blood-brain concordance for the top 10 CpGs sites from the epigenome-wide association study of cerebrospinal fluid-based (CSF) biomarkers of Alzheimer’s disease in the Emory Healthy Brain Study (EHBS, N=450).** | | | | | | | |
| --- | --- | --- | --- | --- | --- | --- | --- |
| **A. tTau** | | | | | | | |
|  |  |  | **BECon** | | | **Gene Expression Omnibus Database** | |
| **CpG** | **Position** | **Gene** | **Mean Cor All Brain** | **Percentile of Mean Cor All Brain (positive)** | **Percentile of Mean Cor All Brain (negative)** | **Brain-blood correlation** | **p-value** |
| cg25530374 | chr16:2047171 | AC005606.15,ZNF598 | n/a | n/a | n/a | 0.064 | 0.784 |
| cg03586820 | chr1:16679780 | SZRD1 | 0.017 | <50% | - | 0.069 | 0.767 |
| cg03376719 | chr3:105086940 | ALCAM | 0.173 | 50-75% | - | -0.222 | 0.332 |
| cg19196826 | chr7:3018391 | CARD11 | 0.067 | <50% | - | -0.425 | 0.056 |
| cg19769827 | chr1:203259772 | RP11-134P9.3 | -0.155 | - | 50-75% | -0.144 | 0.531 |
| cg21719937 | chr12:5564478 | NTF3 | 0.156 | 50-75% | - | -0.327 | 0.148 |
| cg09766383 | chr6:97285174 | GPR63 | -0.025 | - | <50% | 0.066 | 0.776 |
| cg16602332 | chr2:26735409 | OTOF | n/a | n/a | n/a | 0.500 | 0.022 |
| cg06334093 | chr6:139094587 | CCDC28A | -0.010 | - | <50% | -0.084 | 0.716 |
| cg22546737 | chr10:118934495 | RP11-501J20.2 | -0.095 | - | <50% | 0.462 | 0.036 |
| **B. pTau** | | | | | | | |
|  |  |  | **BECon** | | | **Gene Expression Omnibus Database** | |
| **CpG** | **Position** | **Gene** | **Mean Cor All Brain** | **Percentile of Mean Cor All Brain (positive)** | **Percentile of Mean Cor All Brain (negative)** | **Brain-blood correlation** | **p-value** |
| cg03376719 | chr3:105086940 | ALCAM | 0.173 | 50-75% | - | -0.222 | 0.332 |
| cg19196826 | chr7:3018391 | CARD11 | 0.067 | <50% | - | -0.425 | 0.056 |
| cg25530374 | chr16:2047171 | AC005606.15,ZNF598 | n/a | n/a | n/a | 0.064 | 0.784 |
| cg06334093 | chr6:139094587 | CCDC28A | -0.010 | - | <50% | -0.084 | 0.716 |
| cg03586820 | chr1:16679780 | SZRD1 | 0.017 | <50% | - | 0.069 | 0.767 |
| cg13422045 | chr11:73021272 | ARHGEF17 | n/a | n/a | n/a | 0.081 | 0.728 |
| cg09766383 | chr6:97285174 | GPR63 | -0.025 | - | <50% | 0.066 | 0.776 |
| cg21719937 | chr12:5564478 | NTF3 | 0.156 | 50-75% | - | -0.327 | 0.148 |
| cg08186837 | chr1:117910444 | MAN1A2 | n/a | n/a | n/a | 0.230 | 0.315 |
| cg19769827 | chr1:203259772 | RP11-134P9.3 | -0.155 | - | 50-75% | -0.144 | 0.531 |
| **C. Aβ42/tTau** | | | | | | | |
|  |  |  | **BECon** | | | **Gene Expression Omnibus Database** | |
| **CpG** | **Position** | **Gene** | **Mean Cor All Brain** | **Percentile of Mean Cor All Brain (positive)** | **Percentile of Mean Cor All Brain (negative)** | **Brain-blood correlation** | **p-value** |
| cg03586820 | chr1:16679780 | SZRD1 | 0.017 | <50% | - | 0.069 | 0.767 |
| cg10917153 | chr15:42448786 | PLA2G4F | -0.257 | - | 75-90% | 0.083 | 0.720 |
| cg13974715 | chr1:236009306 | LYST | n/a | n/a | n/a | 0.139 | 0.547 |
| cg09340250 | chr1:152924562 | RP1-13P20.6 | n/a | n/a | n/a | 0.206 | 0.367 |
| cg11069276 | chr1:11718175 | FBXO44 | 0.099 | <50% | - | 0.087 | 0.707 |
| cg10235683 | chr8:142304416 | SLC45A4 | n/a | n/a | n/a | 0.179 | 0.435 |
| cg00056692 | chr10:134947537 | GPR123 | n/a | n/a | n/a | 0.244 | 0.285 |
| cg16158487 | chr1:33891592 | PHC2 | n/a | n/a | n/a | -0.001 | 0.998 |
| cg27030540 | chr19:41754975 | AXL | n/a | n/a | n/a | 0.142 | 0.539 |
| cg05832751 | chr8:49716954 | EFCAB1 | n/a | n/a | n/a | -0.299 | 0.188 |
| **D. Aβ42+/-** | | | | | | | |
|  |  |  | **BECon** | | | **Gene Expression Omnibus Database** | |
| **CpG** | **Position** | **Gene** | **Mean Cor All Brain** | **Percentile of Mean Cor All Brain (positive)** | **Percentile of Mean Cor All Brain (negative)** | **Brain-blood correlation** | **p-value** |
| cg08759359 | chr5:3288934 | CTD-2029E14.1 | -0.060 | - | <50% | -0.203 | 0.377 |
| cg27047965 | chr1:54433172 | LRRC42 | n/a | n/a | n/a | -0.174 | 0.449 |
| cg15226147 | chr19:1275266 | C19orf24 | -0.021 | - | <50% | -0.195 | 0.396 |
| cg27504433 | chr7:6741096 | ZNF12 | n/a | n/a | n/a | -0.016 | 0.948 |
| cg06769708 | chr20:35060706 | DLGAP4 | -0.162 | - | 50-75% | -0.191 | 0.405 |
| cg18890561 | chr10:131988419 | GLRX3 | -0.008 | - | <50% | 0.100 | 0.665 |
| cg20673767 | chr7:158061805 | PTPRN2 | 0.017 | <50% | - | 0.316 | 0.163 |
| cg20751395 | chr11:2594153 | KCNQ1 | -0.225 | - | 50-75% | -0.082 | 0.724 |
| cg16879549 | chr17:7146439 | CTD-2545G14.7 | 0.451 | 90% | - | 0.636 | 0.002 |
| cg02400458 | chr15:80624605 | LINC00927 | -0.054 | - | <50% | 0.074 | 0.750 |
| **E. pTau+/-** |  | | | | | | |
|  |  |  | **BECon** | | | **Gene Expression Omnibus Database** | |
| **CpG** | **Position** | **Gene** | **Mean Cor All Brain** | **Percentile of Mean Cor All Brain (positive)** | **Percentile of Mean Cor All Brain (negative)** | **Brain-blood correlation** | **p-value** |
| cg18254930 | chr3:3646624 | AC026188.1 | -0.229 | - | 75-90% | 0.378 | 0.092 |
| cg06763914 | chr16:74260395 | AC009120.4 | n/a | n/a | n/a | 0.281 | 0.217 |
| cg11175683 | chr7:94286420 | PEG10 | 0.040 | <50% | - | -0.058 | 0.802 |
| cg16348003 | chr1:153589781 | S100A14 | 0.306 | 75-90% | - | 0.418 | 0.060 |
| cg13422045 | chr11:73021272 | ARHGEF17 | n/a | n/a | n/a | 0.081 | 0.728 |
| cg15104031 | chr1:153589528 | S100A14 | 0.205 | 50-75% | - | 0.253 | 0.267 |
| cg01928691 | chr2:106016014 | FHL2 | n/a | n/a | n/a | 0.314 | 0.165 |
| cg21719937 | chr12:5564478 | NTF3 | 0.156 | 50-75% | - | -0.327 | 0.148 |
| cg05734494 | chr17:57287309 | SMG8 | n/a | n/a | n/a | 0.216 | 0.346 |
| cg11537121 | chr4:184575108 | RWDD4 | n/a | n/a | n/a | 0.338 | 0.135 |

| **Table S4. Blood-brain concordance for the top 10 CpGs sites from the epigenome-wide association study of cerebrospinal fluid-based (CSF) biomarkers of Alzheimer’s disease in ADNI (N=122)** | | | | | | | |
| --- | --- | --- | --- | --- | --- | --- | --- |
| **A. tTau** | | | | | | | |
|  |  |  | **BECon** | | | **Gene Expression Omnibus Database** | |
| **CpG** | **Position** | **Gene** | **Mean Cor All Brain** | **Percentile of Mean Cor All Brain (positive)** | **Percentile of Mean Cor All Brain (negative)** | **Brain-blood correlation** | **p-value** |
| cg08522625 | chr3:46135532 | FLT1P1 | n/a | n/a | n/a | -0.284 | 0.211 |
| cg10071605 | chr16:31072718 | ZNF668 | n/a | n/a | n/a | 0.255 | 0.264 |
| cg17162734 | chr10:71337586 | RP11-343J3.2 | -0.116 | - | <50% | 0.344 | 0.127 |
| cg13468451 | chr14:101315775 | MEG3 | n/a | n/a | n/a | 0.008 | 0.975 |
| cg12833422 | chr11:118166049 | CD3E | n/a | n/a | n/a | 0.606 | 0.004 |
| cg19026692 | chr2:168947421 | STK39 | n/a | n/a | n/a | 0.300 | 0.186 |
| cg18581037 | chr5:169010289 | SPDL1 | -0.358 | - | 90% | 0.330 | 0.144 |
| cg10604715 | chr4:2279733 | ZFYVE28 | n/a | n/a | n/a | 0.196 | 0.393 |
| cg22868272 | chr8:65936215 | RP11-89A16.1 | -0.330 | - | 90% | -0.217 | 0.343 |
| cg19518069 | chr12:10320770 | OLR1 | n/a | n/a | n/a | -0.470 | 0.033 |
| **B. pTau** | | | | | | | |
|  |  |  | **BECon** | | | **Gene Expression Omnibus Database** | |
| **CpG** | **Position** | **Gene** | **Mean Cor All Brain** | **Percentile of Mean Cor All Brain (positive)** | **Percentile of Mean Cor All Brain (negative)** | **Brain-blood correlation** | **p-value** |
| cg21416544 | chr12:8986988 | A2ML1 | -0.182 | - | 50-75% | -0.075 | 0.745 |
| cg10071605 | chr16:31072718 | ZNF668 | n/a | n/a | n/a | 0.255 | 0.264 |
| cg13468451 | chr14:101315775 | MEG3 | n/a | n/a | n/a | 0.008 | 0.975 |
| cg19026692 | chr2:168947421 | STK39 | n/a | n/a | n/a | 0.300 | 0.186 |
| cg12833422 | chr11:118166049 | CD3E | n/a | n/a | n/a | 0.606 | 0.004 |
| cg08522625 | chr3:46135532 | FLT1P1 | n/a | n/a | n/a | -0.284 | 0.211 |
| cg17162734 | chr10:71337586 | RP11-343J3.2 | -0.116 | - | <50% | 0.344 | 0.127 |
| cg18581037 | chr5:169010289 | SPDL1 | -0.358 | - | 90% | 0.330 | 0.144 |
| cg11910223 | chr5:139060821 | CXXC5 | -0.059 | - | <50% | -0.038 | 0.872 |
| cg22868272 | chr8:65936215 | RP11-89A16.1 | -0.330 | - | 90% | -0.217 | 0.343 |
| **C. Aβ42/tTau** | | | | | | | |
|  |  |  | **BECon** | | | **Gene Expression Omnibus Database** | |
| **CpG** | **Position** | **Gene** | **Mean Cor All Brain** | **Percentile of Mean Cor All Brain (positive)** | **Percentile of Mean Cor All Brain (negative)** | **Brain-blood correlation** | **p-value** |
| cg21021972 | chr3:64613818 | ADAMTS9 | n/a | n/a | n/a | -0.260 | 0.254 |
| cg04434425 | chr6:22929545 | RP1-209A6.1 | n/a | n/a | n/a | 0.108 | 0.641 |
| cg09034302 | chr7:95108238 | ASB4 | n/a | n/a | n/a | 0.229 | 0.317 |
| cg01028017 | chr6:92363854 | CASC6 | -0.163 | - | 50-75% | -0.101 | 0.661 |
| cg04985396 | chr1:72749735 | NEGR1 | -0.284 | - | 75-90% | 0.304 | 0.180 |
| cg16729690 | chr8:28351217 | FZD3 | -0.067 | - | <50% | 0.043 | 0.854 |
| cg03455845 | chr1:96840865 | RP5-898J17.1 | n/a | n/a | n/a | 0.121 | 0.601 |
| cg27234746 | chr5:170579510 | RANBP17 | n/a | n/a | n/a | 0.097 | 0.674 |
| cg24738424 | chr6:169364882 | RP3-495K2.3 | n/a | n/a | n/a | 0.039 | 0.868 |
| cg10037002 | chr20:36378118 | CTNNBL1 | n/a | n/a | n/a | -0.105 | 0.649 |
| **D. Aβ42+/-** | | | | | | | |
|  |  |  | **BECon** | | | **Gene Expression Omnibus Database** | |
| **CpG** | **Position** | **Gene** | **Mean Cor All Brain** | **Percentile of Mean Cor All Brain (positive)** | **Percentile of Mean Cor All Brain (negative)** | **Brain-blood correlation** | **p-value** |
| cg17394124 | chr1:196712668 | CFH | n/a | n/a | n/a | -0.222 | 0.332 |
| cg08770004 | chr12:125170382 | SCARB1 | n/a | n/a | n/a | -0.231 | 0.312 |
| cg17394795 | chr9:96628794 | RP11-53B5.1 | n/a | n/a | n/a | 0.068 | 0.771 |
| cg04903883 | chr14:106321551 | IGHM | -0.424 | - | 90% | 0.106 | 0.645 |
| cg06014227 | chr12:13028038 | RPL13AP20 | -0.081 | - | <50% | -0.030 | 0.899 |
| cg14123436 | chr9:98544579 | LINC00476 | -0.088 | - | <50% | 0.081 | 0.728 |
| cg06869529 | chr8:94594936 | LINC00535 | n/a | n/a | n/a | 0.227 | 0.320 |
| cg16624991 | chr8:119931917 | TNFRSF11B | n/a | n/a | n/a | 0.005 | 0.984 |
| cg12252090 | chr8:49833279 | SNAI2 | -0.228 | - | 75-90% | 0.235 | 0.304 |
| cg17394124 | chr1:196712668 | CFH | n/a | n/a | n/a | -0.222 | 0.332 |
| **E. pTau+/-** |  | | | | | | |
|  |  |  | **BECon** | | | **Gene Expression Omnibus Database** | |
| **CpG** | **Position** | **Gene** | **Mean Cor All Brain** | **Percentile of Mean Cor All Brain (positive)** | **Percentile of Mean Cor All Brain (negative)** | **Brain-blood correlation** | **p-value** |
| cg07487535 | chr11:71159905 | RP11-660L16.2 | -0.211 | - | 50-75% | 0.139 | 0.547 |
| cg16666973 | chr12:64492812 | SRGAP1 | n/a | n/a | n/a | 0.168 | 0.466 |
| cg06233376 | chr16:6532764 | RBFOX1 | -0.166 | - | 50-75% | 0.084 | 0.716 |
| cg15253520 | chr8:87521369 | RMDN1 | 0.286 | 75-90% | - | 0.019 | 0.935 |
| cg17104202 | chr1:3162404 | PRDM16 | -0.333 | - | 90% | -0.149 | 0.517 |
| cg05853285 | chr11:44339621 | ALX4 | -0.036 | - | <50% | 0.378 | 0.092 |
| cg00879843 | chr6:33239764 | VPS52 | -0.275 | - | 75-90% | 0.122 | 0.597 |
| cg14389924 | chr17:4606729 | PELP1 | 0.052 | <50% | - | -0.153 | 0.506 |
| cg00611605 | chr6:107236467 | RP1-60O19.1 | n/a | n/a | n/a | 0.208 | 0.364 |
| cg03459510 | chr12:114300370 | RBM19 | n/a | n/a | n/a | 0.188 | 0.412 |

| **Table S5. Blood-brain concordance for the top 10 CpGs sites from the epigenome-wide meta-analysis of cerebrospinal fluid-based (CSF) biomarkers of Alzheimer’s disease in in 450 cognitively normal individuals from the Emory Healthy Brain Study (EHBS) and 122 cognitively normal individuals from ADNI.** | | | | | | | |
| --- | --- | --- | --- | --- | --- | --- | --- |
| **A. tTau** | | | | | | | |
|  |  |  | **BECon** | | | **Gene Expression Omnibus Database** | |
| **CpG** | **Position** | **Gene** | **Mean Cor All Brain** | **Percentile of Mean Cor All Brain (positive)** | **Percentile of Mean Cor All Brain (negative)** | **Brain-blood correlation** | **p-value** |
| cg25530374 | chr16:2047171 | AC005606.15,ZNF598 | n/a | n/a | n/a | 0.064 | 0.784 |
| cg03586820 | chr1:16679780 | SZRD1 | 0.017 | <50% | - | 0.069 | 0.767 |
| cg19196826 | chr7:3018391 | CARD11 | 0.067 | <50% | - | -0.425 | 0.056 |
| cg03376719 | chr3:105086940 | ALCAM | 0.173 | 50-75% | - | -0.222 | 0.332 |
| cg19769827 | chr1:203259772 | RP11-134P9.3 | -0.155 | - | 50-75% | -0.144 | 0.531 |
| cg21719937 | chr12:5564478 | NTF3 | 0.156 | 50-75% | - | -0.327 | 0.148 |
| cg22546737 | chr10:118934495 | RP11-501J20.2 | -0.095 | - | <50% | 0.462 | 0.036 |
| cg05104523 | chr4:3295914 | RGS12 | n/a | n/a | n/a | 0.535 | 0.014 |
| cg16602332 | chr2:26735409 | OTOF | n/a | n/a | n/a | 0.500 | 0.022 |
| cg09766383 | chr6:97285174 | GPR63 | -0.025 | - | <50% | 0.066 | 0.776 |
| **B. pTau** | | | | | | | |
|  |  |  | **BECon** | | | **Gene Expression Omnibus Database** | |
| **CpG** | **Position** | **Gene** | **Mean Cor All Brain** | **Percentile of Mean Cor All Brain (positive)** | **Percentile of Mean Cor All Brain (negative)** | **Brain-blood correlation** | **p-value** |
| cg25530374 | chr16:2047171 | AC005606.15,ZNF598 | n/a | n/a | n/a | 0.064 | 0.784 |
| cg19196826 | chr7:3018391 | CARD11 | 0.067 | <50% | - | -0.425 | 0.056 |
| cg03376719 | chr3:105086940 | ALCAM | 0.173 | 50-75% | - | -0.222 | 0.332 |
| cg05104523 | chr4:3295914 | RGS12 | n/a | n/a | n/a | 0.535 | 0.014 |
| cg06334093 | chr6:139094587 | CCDC28A | -0.010 | - | <50% | -0.084 | 0.716 |
| cg03586820 | chr1:16679780 | SZRD1 | 0.017 | <50% | - | 0.069 | 0.767 |
| cg22546737 | chr10:118934495 | RP11-501J20.2 | -0.095 | - | <50% | 0.462 | 0.036 |
| cg00021892 | chr1:153582521 | S100A16 | n/a | n/a | n/a | 0.283 | 0.213 |
| cg25377744 | chr13:47063673 | COX17P1 | -0.323 | - | 90% | -0.395 | 0.078 |
| cg21719937 | chr12:5564478 | NTF3 | 0.156 | 50-75% | - | -0.327 | 0.148 |
| **C. Aβ42/tTau** | | | | | | | |
|  |  |  | **BECon** | | | **Gene Expression Omnibus Database** | |
| **CpG** | **Position** | **Gene** | **Mean Cor All Brain** | **Percentile of Mean Cor All Brain (positive)** | **Percentile of Mean Cor All Brain (negative)** | **Brain-blood correlation** | **p-value** |
| cg03586820 | chr1:16679780 | SZRD1 | 0.017 | <50% | - | 0.069 | 0.767 |
| cg10917153 | chr15:42448786 | PLA2G4F | -0.257 | - | 75-90% | 0.083 | 0.720 |
| cg13974715 | chr1:236009306 | LYST | n/a | n/a | n/a | 0.139 | 0.547 |
| cg09340250 | chr1:152924562 | RP1-13P20.6 | n/a | n/a | n/a | 0.206 | 0.367 |
| cg07311033 | chr2:111627862 | ACOXL | n/a | n/a | n/a | 0.231 | 0.312 |
| cg25381285 | chr14:102691354 | MOK | n/a | n/a | n/a | 0.143 | 0.535 |
| cg10633103 | chr20:43561125 | PABPC1L | n/a | n/a | n/a | 0.378 | 0.092 |
| cg10235683 | chr8:142304416 | SLC45A4 | n/a | n/a | n/a | 0.179 | 0.435 |
| cg14933468 | chr11:62138599 | ASRGL1 | 0.076 | <50% | - | 0.142 | 0.539 |
| cg16158487 | chr1:33891592 | PHC2 | n/a | n/a | n/a | -0.001 | 0.998 |
| **D. Aβ42+/-** | | | | | | | |
|  |  |  | **BECon** | | | **Gene Expression Omnibus Database** | |
| **CpG** | **Position** | **Gene** | **Mean Cor All Brain** | **Percentile of Mean Cor All Brain (positive)** | **Percentile of Mean Cor All Brain (negative)** | **Brain-blood correlation** | **p-value** |
| cg08216368 | chr11:237063 | PSMD13 | -0.320 | - | 90% | 0.140 | 0.543 |
| cg17394795 | chr9:96628794 | RP11-53B5.1 | n/a | n/a | n/a | 0.068 | 0.771 |
| cg18890561 | chr10:131988419 | GLRX3 | -0.008 | - | <50% | 0.100 | 0.665 |
| cg27504433 | chr7:6741096 | ZNF12 | n/a | n/a | n/a | -0.016 | 0.948 |
| cg18407095 | chr2:26846581 | CIB4 | -0.386 | - | 90% | 0.119 | 0.605 |
| cg13589108 | chr1:177140680 | BRINP2 | -0.089 | - | <50% | -0.418 | 0.060 |
| cg05961166 | chr21:26864304 | snoU13 | n/a | n/a | n/a | 0.284 | 0.211 |
| cg16182707 | chr12:105478090 | ALDH1L2 | -0.072 | - | <50% | 0.169 | 0.463 |
| cg07767421 | chr16:5059086 | SEC14L5 | n/a | n/a | n/a | 0.045 | 0.846 |
| cg06769708 | chr20:35060706 | DLGAP4 | -0.162 | - | 50-75% | -0.191 | 0.405 |
| **E. pTau+/-** |  | | | | | | |
|  |  |  | **BECon** | | | **Gene Expression Omnibus Database** | |
| **CpG** | **Position** | **Gene** | **Mean Cor All Brain** | **Percentile of Mean Cor All Brain (positive)** | **Percentile of Mean Cor All Brain (negative)** | **Brain-blood correlation** | **p-value** |
| cg01928691 | chr2:106016014 | FHL2 | n/a | n/a | n/a | 0.314 | 0.165 |
| cg22207257 | chr1:193361991 | LINC01031 | n/a | n/a | n/a | 0.299 | 0.188 |
| cg18254930 | chr3:3646624 | AC026188.1 | -0.229 | - | 75-90% | 0.378 | 0.092 |
| cg12031108 | chr12:28115086 | PTHLH | n/a | n/a | n/a | 0.166 | 0.470 |
| cg16348003 | chr1:153589781 | S100A14 | 0.306 | 75-90% | - | 0.418 | 0.060 |
| cg18958053 | chr15:55348892 | RP11-548M13.1 | n/a | n/a | n/a | 0.310 | 0.171 |
| cg02671700 | chr8:64523255 | RN7SKP135 | n/a | n/a | n/a | 0.100 | 0.665 |
| cg16882206 | chr8:115466108 | RP11-393K19.1 | 0.169 | 50-75% | - | 0.013 | 0.957 |
| cg10935297 | chr14:35255491 | BAZ1A | n/a | n/a | n/a | -0.135 | 0.558 |
| cg02777461 | chr13:63801462 | LINC00376 | n/a | n/a | n/a | -0.031 | 0.895 |

**Table S6. First 10 GO terms with raw pvalue < 0.05 from the epigenome-wide association study of cerebrospinal fluid-based (CSF) biomarkers of Alzheimer’s disease in the Emory Healthy Brain Study (EHBS, N=450).**

| **A. tTau** | | | | | |
| --- | --- | --- | --- | --- | --- |
| **GO term** | **ONTOLOGY** | **TERM** | **N** | **DE** | **P.DE** |
| GO:0033260 | BP | nuclear DNA replication | 39 | 8 | 2.87E-04 |
| GO:0044786 | BP | cell cycle DNA replication | 43 | 8 | 5.50E-04 |
| GO:0032201 | BP | telomere maintenance via semi-conservative replication | 9 | 4 | 5.97E-04 |
| GO:0016055 | BP | Wnt signaling pathway | 444 | 39 | 1.04E-03 |
| GO:0198738 | BP | cell-cell signaling by wnt | 446 | 39 | 1.15E-03 |
| GO:1905912 | BP | regulation of calcium ion export across plasma membrane | 6 | 3 | 1.25E-03 |
| GO:0042162 | MF | telomeric DNA binding | 37 | 7 | 1.35E-03 |
| GO:0031929 | BP | TOR signaling | 155 | 17 | 1.43E-03 |
| GO:0003720 | MF | telomerase activity | 6 | 3 | 2.11E-03 |
| GO:0032213 | BP | regulation of telomere maintenance via semi-conservative replication | 2 | 2 | 2.57E-03 |
| **B. pTau** | | | | | |
| **GO term** | **ONTOLOGY** | **TERM** | **N** | **DE** | **P.DE** |
| GO:0003401 | BP | axis elongation | 28 | 7 | 7.55E-04 |
| GO:0051386 | BP | regulation of neurotrophin TRK receptor signaling pathway | 10 | 4 | 1.20E-03 |
| GO:0010921 | BP | regulation of phosphatase activity | 57 | 9 | 1.27E-03 |
| GO:0043666 | BP | regulation of phosphoprotein phosphatase activity | 45 | 8 | 1.36E-03 |
| GO:1904016 | BP | response to Thyroglobulin triiodothyronine | 5 | 3 | 2.02E-03 |
| GO:0030011 | BP | maintenance of cell polarity | 17 | 5 | 2.09E-03 |
| GO:0003720 | MF | telomerase activity | 6 | 3 | 2.24E-03 |
| GO:0006470 | BP | protein dephosphorylation | 201 | 21 | 2.33E-03 |
| GO:0060560 | BP | developmental growth involved in morphogenesis | 230 | 25 | 2.52E-03 |
| GO:0032213 | BP | regulation of telomere maintenance via semi-conservative replication | 2 | 2 | 2.63E-03 |
| **C. Aβ42/tTau** | | | | | |
| **GO term** | **ONTOLOGY** | **TERM** | **N** | **DE** | **P.DE** |
| GO:0072079 | BP | nephron tubule formation | 20 | 6 | 2.05E-04 |
| GO:0009052 | BP | pentose-phosphate shunt, non-oxidative branch | 6 | 3 | 6.04E-04 |
| GO:0043266 | BP | regulation of potassium ion transport | 99 | 13 | 7.14E-04 |
| GO:0072106 | BP | regulation of ureteric bud formation | 4 | 3 | 8.04E-04 |
| GO:0072107 | BP | positive regulation of ureteric bud formation | 4 | 3 | 8.04E-04 |
| GO:0005225 | MF | volume-sensitive anion channel activity | 9 | 4 | 1.14E-03 |
| GO:0006098 | BP | pentose-phosphate shunt | 19 | 5 | 1.16E-03 |
| GO:0051156 | BP | glucose 6-phosphate metabolic process | 28 | 6 | 1.25E-03 |
| GO:1901379 | BP | regulation of potassium ion transmembrane transport | 83 | 11 | 1.57E-03 |
| GO:0046651 | BP | lymphocyte proliferation | 288 | 22 | 1.78E-03 |
| **D. Aβ42+/-** | | | | | |
| **GO term** | **ONTOLOGY** | **TERM** | **N** | **DE** | **P.DE** |
| GO:0002415 | BP | immunoglobulin transcytosis in epithelial cells mediated by polymeric immunoglobulin receptor | 2 | 2 | 7.78E-04 |
| GO:0033233 | BP | regulation of protein sumoylation | 21 | 5 | 1.47E-03 |
| GO:0061436 | BP | establishment of skin barrier | 33 | 6 | 1.48E-03 |
| GO:0016492 | MF | G protein-coupled neurotensin receptor activity | 2 | 2 | 1.79E-03 |
| GO:0071864 | BP | positive regulation of cell proliferation in bone marrow | 6 | 3 | 2.28E-03 |
| GO:1901607 | BP | alpha-amino acid biosynthetic process | 65 | 8 | 2.40E-03 |
| GO:0004084 | MF | branched-chain-amino-acid transaminase activity | 2 | 2 | 2.86E-03 |
| GO:0009098 | BP | leucine biosynthetic process | 2 | 2 | 2.86E-03 |
| GO:0050048 | MF | L-leucine:2-oxoglutarate aminotransferase activity | 2 | 2 | 2.86E-03 |
| GO:0052654 | MF | L-leucine transaminase activity | 2 | 2 | 2.86E-03 |
| **E. pTau+/-** | | | | | |
| **GO term** | **ONTOLOGY** | **TERM** | **N** | **DE** | **P.DE** |
| GO:0090201 | BP | negative regulation of release of cytochrome c from mitochondria | 19 | 6 | 5.15E-05 |
| GO:0070374 | BP | positive regulation of ERK1 and ERK2 cascade | 212 | 20 | 1.14E-04 |
| GO:0042660 | BP | positive regulation of cell fate specification | 3 | 3 | 2.43E-04 |
| GO:0003713 | MF | transcription coactivator activity | 257 | 25 | 3.64E-04 |
| GO:0097528 | BP | execution phase of necroptosis | 2 | 2 | 6.60E-04 |
| GO:0141124 | BP | intracellular signaling cassette | 1822 | 107 | 6.73E-04 |
| GO:0008631 | BP | intrinsic apoptotic signaling pathway in response to oxidative stress | 62 | 9 | 8.05E-04 |
| GO:0060978 | BP | angiogenesis involved in coronary vascular morphogenesis | 5 | 3 | 9.04E-04 |
| GO:0019369 | BP | arachidonic acid metabolic process | 55 | 7 | 1.21E-03 |
| GO:0070372 | BP | regulation of ERK1 and ERK2 cascade | 299 | 23 | 1.33E-03 |

**Table S7. First 10 GO terms with raw pvalue < 0.05 from the epigenome-wide association study of cerebrospinal fluid-based (CSF) biomarkers of Alzheimer’s disease in ADNI (N=122).**

| **A. tTau** | | | | | |
| --- | --- | --- | --- | --- | --- |
| **GO term** | **ONTOLOGY** | **TERM** | **N** | **DE** | **P.DE** |
| GO:0034685 | CC | integrin alphav-beta6 complex | 2 | 2 | 9.12E-04 |
| GO:0035697 | BP | CD8-positive, alpha-beta T cell extravasation | 3 | 2 | 1.46E-03 |
| GO:2000449 | BP | regulation of CD8-positive, alpha-beta T cell extravasation | 3 | 2 | 1.46E-03 |
| GO:0035579 | CC | specific granule membrane | 88 | 10 | 2.73E-03 |
| GO:0046332 | MF | SMAD binding | 74 | 11 | 2.77E-03 |
| GO:0016312 | MF | inositol bisphosphate phosphatase activity | 6 | 3 | 3.19E-03 |
| GO:0010815 | BP | bradykinin catabolic process | 7 | 3 | 3.64E-03 |
| GO:0030613 | MF | oxidoreductase activity, acting on phosphorus or arsenic in donors | 3 | 2 | 3.85E-03 |
| GO:0030614 | MF | oxidoreductase activity, acting on phosphorus or arsenic in donors, disulfide as acceptor | 3 | 2 | 3.85E-03 |
| GO:0008037 | BP | cell recognition | 153 | 14 | 4.87E-03 |
| **B. pTau** | | | | | |
| **GO term** | **ONTOLOGY** | **TERM** | **N** | **DE** | **P.DE** |
| GO:0005160 | MF | transforming growth factor beta receptor binding | 26 | 7 | 1.02E-04 |
| GO:1903860 | BP | negative regulation of dendrite extension | 3 | 3 | 1.93E-04 |
| GO:0030512 | BP | negative regulation of transforming growth factor beta receptor signaling pathway | 121 | 13 | 5.93E-04 |
| GO:0005114 | MF | type II transforming growth factor beta receptor binding | 11 | 4 | 6.06E-04 |
| GO:0046332 | MF | SMAD binding | 74 | 12 | 6.27E-04 |
| GO:0034685 | CC | integrin alphav-beta6 complex | 2 | 2 | 8.68E-04 |
| GO:0017015 | BP | regulation of transforming growth factor beta receptor signaling pathway | 170 | 16 | 1.37E-03 |
| GO:1903844 | BP | regulation of cellular response to transforming growth factor beta stimulus | 173 | 16 | 1.73E-03 |
| GO:0043124 | BP | negative regulation of canonical NF-kappaB signal transduction | 61 | 8 | 2.30E-03 |
| GO:0060395 | BP | SMAD protein signal transduction | 94 | 11 | 2.51E-03 |
| **C. Aβ42/tTau** | | | | | |
| **GO term** | **ONTOLOGY** | **TERM** | **N** | **DE** | **P.DE** |
| GO:0030900 | BP | forebrain development | 392 | 38 | 3.55E-05 |
| GO:0060322 | BP | head development | 735 | 59 | 6.67E-05 |
| GO:0007420 | BP | brain development | 688 | 56 | 6.91E-05 |
| GO:0004918 | MF | interleukin-8 receptor activity | 2 | 2 | 2.96E-04 |
| GO:0038112 | BP | interleukin-8-mediated signaling pathway | 2 | 2 | 2.96E-04 |
| GO:0048699 | BP | generation of neurons | 1443 | 99 | 3.19E-04 |
| GO:0007409 | BP | axonogenesis | 429 | 41 | 3.63E-04 |
| GO:0019935 | BP | cyclic-nucleotide-mediated signaling | 97 | 13 | 3.82E-04 |
| GO:0030182 | BP | neuron differentiation | 1365 | 94 | 3.97E-04 |
| GO:0090596 | BP | sensory organ morphogenesis | 273 | 26 | 4.72E-04 |
| **D. Aβ42+/-** | | | | | |
| **GO term** | **ONTOLOGY** | **TERM** | **N** | **DE** | **P.DE** |
| GO:0140092 | CC | bBAF complex | 10 | 4 | 1.02E-03 |
| GO:0046398 | BP | UDP-glucuronate metabolic process | 2 | 2 | 2.87E-03 |
| GO:0016191 | BP | synaptic vesicle uncoating | 6 | 3 | 3.28E-03 |
| GO:0043218 | CC | compact myelin | 14 | 4 | 3.66E-03 |
| GO:0035136 | BP | forelimb morphogenesis | 40 | 7 | 3.85E-03 |
| GO:0071564 | CC | npBAF complex | 14 | 4 | 3.92E-03 |
| GO:1900195 | BP | positive regulation of oocyte maturation | 3 | 2 | 4.00E-03 |
| GO:0018201 | BP | peptidyl-glycine modification | 3 | 2 | 4.39E-03 |
| GO:0045176 | BP | apical protein localization | 14 | 4 | 5.26E-03 |
| GO:0035115 | BP | embryonic forelimb morphogenesis | 33 | 6 | 5.31E-03 |
| **E. pTau+/-** | | | | | |
| **GO term** | **ONTOLOGY** | **TERM** | **N** | **DE** | **P.DE** |
| GO:0002444 | BP | myeloid leukocyte mediated immunity | 104 | 13 | 1.50E-04 |
| GO:0008443 | MF | phosphofructokinase activity | 6 | 4 | 2.58E-04 |
| GO:0070668 | BP | positive regulation of mast cell proliferation | 4 | 3 | 5.90E-04 |
| GO:0004657 | MF | proline dehydrogenase activity | 2 | 2 | 5.96E-04 |
| GO:0005078 | MF | MAP-kinase scaffold activity | 12 | 5 | 8.19E-04 |
| GO:0043067 | BP | regulation of programmed cell death | 1466 | 89 | 8.57E-04 |
| GO:0071889 | MF | 14-3-3 protein binding | 32 | 8 | 9.10E-04 |
| GO:0002573 | BP | myeloid leukocyte differentiation | 230 | 22 | 1.03E-03 |
| GO:0005160 | MF | transforming growth factor beta receptor binding | 26 | 6 | 1.15E-03 |
| GO:0048870 | BP | cell motility | 1740 | 107 | 1.24E-03 |

**Table S8. First 10 GO terms with raw pvalue < 0.05 from the epigenome-wide meta-analysis of cerebrospinal fluid-based (CSF) biomarkers of Alzheimer’s disease in in 450 cognitively normal individuals from the Emory Healthy Brain Study (EHBS) and 122 cognitively normal individuals from ADNI.**

| **A. tTau** | | | | | |
| --- | --- | --- | --- | --- | --- |
| **GO term** | **ONTOLOGY** | **TERM** | **N** | **DE** | **P.DE** |
| GO:0016055 | BP | Wnt signaling pathway | 444 | 40 | 6.15E-04 |
| GO:0198738 | BP | cell-cell signaling by wnt | 446 | 40 | 6.82E-04 |
| GO:0031929 | BP | TOR signaling | 155 | 17 | 1.45E-03 |
| GO:0033260 | BP | nuclear DNA replication | 39 | 7 | 1.56E-03 |
| GO:0044786 | BP | cell cycle DNA replication | 43 | 7 | 2.65E-03 |
| GO:0045577 | BP | regulation of B cell differentiation | 32 | 6 | 2.99E-03 |
| GO:0032398 | CC | MHC class Ib protein complex | 2 | 2 | 3.21E-03 |
| GO:0050291 | MF | sphingosine N-acyltransferase activity | 7 | 3 | 3.24E-03 |
| GO:0009646 | BP | response to absence of light | 2 | 2 | 4.46E-03 |
| GO:0071485 | BP | cellular response to absence of light | 2 | 2 | 4.46E-03 |
| **B. pTau** | | | | | |
| **GO term** | **ONTOLOGY** | **TERM** | **N** | **DE** | **P.DE** |
| GO:0051386 | BP | regulation of neurotrophin TRK receptor signaling pathway | 10 | 5 | 6.95E-05 |
| GO:0035088 | BP | establishment or maintenance of apical/basal cell polarity | 53 | 11 | 1.44E-04 |
| GO:0061245 | BP | establishment or maintenance of bipolar cell polarity | 53 | 11 | 1.44E-04 |
| GO:0016421 | MF | CoA carboxylase activity | 6 | 4 | 2.54E-04 |
| GO:0045935 | BP | positive regulation of nucleobase-containing compound metabolic process | 2034 | 126 | 3.13E-04 |
| GO:0045197 | BP | establishment or maintenance of epithelial cell apical/basal polarity | 49 | 10 | 3.92E-04 |
| GO:0016885 | MF | ligase activity, forming carbon-carbon bonds | 7 | 4 | 6.67E-04 |
| GO:0048538 | BP | thymus development | 49 | 9 | 7.34E-04 |
| GO:0051254 | BP | positive regulation of RNA metabolic process | 1837 | 113 | 9.57E-04 |
| GO:0070373 | BP | negative regulation of ERK1 and ERK2 cascade | 68 | 10 | 1.09E-03 |
| **C. Aβ42/tTau** | | | | | |
| **GO term** | **ONTOLOGY** | **TERM** | **N** | **DE** | **P.DE** |
| GO:0004658 | MF | propionyl-CoA carboxylase activity | 2 | 2 | 4.13E-03 |
| GO:1901379 | BP | regulation of potassium ion transmembrane transport | 83 | 10 | 4.47E-03 |
| GO:0006971 | BP | hypotonic response | 13 | 4 | 5.06E-03 |
| GO:0005589 | CC | collagen type VI trimer | 2 | 2 | 5.13E-03 |
| GO:0098647 | CC | collagen beaded filament | 2 | 2 | 5.13E-03 |
| GO:0003976 | MF | UDP-N-acetylglucosamine-lysosomal-enzyme N-acetylglucosaminephosphotransferase activity | 3 | 2 | 5.14E-03 |
| GO:0043376 | BP | regulation of CD8-positive, alpha-beta T cell differentiation | 8 | 3 | 6.03E-03 |
| GO:0051156 | BP | glucose 6-phosphate metabolic process | 28 | 5 | 6.69E-03 |
| GO:1903596 | BP | regulation of gap junction assembly | 9 | 3 | 7.13E-03 |
| GO:0032483 | BP | regulation of Rab protein signal transduction | 7 | 3 | 7.62E-03 |
| **D. Aβ42+/-** | | | | | |
| **GO term** | **ONTOLOGY** | **TERM** | **N** | **DE** | **P.DE** |
| GO:0010801 | BP | negative regulation of peptidyl-threonine phosphorylation | 19 | 5 | 8.24E-04 |
| GO:0018210 | BP | peptidyl-threonine modification | 103 | 13 | 1.86E-03 |
| GO:0018107 | BP | peptidyl-threonine phosphorylation | 93 | 12 | 2.15E-03 |
| GO:0010799 | BP | regulation of peptidyl-threonine phosphorylation | 42 | 7 | 2.24E-03 |
| GO:0004776 | MF | succinate-CoA ligase (GDP-forming) activity | 2 | 2 | 2.28E-03 |
| GO:0045244 | CC | succinate-CoA ligase complex (GDP-forming) | 2 | 2 | 2.28E-03 |
| GO:0003348 | BP | cardiac endothelial cell differentiation | 6 | 3 | 3.19E-03 |
| GO:0060956 | BP | endocardial cell differentiation | 6 | 3 | 3.19E-03 |
| GO:0016209 | MF | antioxidant activity | 83 | 8 | 3.21E-03 |
| GO:0060316 | BP | positive regulation of ryanodine-sensitive calcium-release channel activity | 8 | 3 | 3.27E-03 |
| **E. pTau+/-** | | | | | |
| **GO term** | **ONTOLOGY** | **TERM** | **N** | **DE** | **P.DE** |
| GO:0016942 | CC | insulin-like growth factor binding protein complex | 5 | 3 | 4.29E-04 |
| GO:0097528 | BP | execution phase of necroptosis | 2 | 2 | 4.86E-04 |
| GO:0160006 | BP | Fc receptor-mediated immune complex endocytosis | 2 | 2 | 5.73E-04 |
| GO:0003713 | MF | transcription coactivator activity | 257 | 22 | 1.29E-03 |
| GO:2001187 | BP | positive regulation of CD8-positive, alpha-beta T cell activation | 7 | 3 | 2.39E-03 |
| GO:0005031 | MF | tumor necrosis factor receptor activity | 8 | 3 | 2.70E-03 |
| GO:0051155 | BP | positive regulation of striated muscle cell differentiation | 52 | 7 | 2.78E-03 |
| GO:0051000 | BP | positive regulation of nitric-oxide synthase activity | 15 | 4 | 2.81E-03 |
| GO:0002692 | BP | negative regulation of cellular extravasation | 9 | 3 | 2.97E-03 |
| GO:0036454 | CC | growth factor complex | 8 | 3 | 3.16E-03 |

**Table S9. First 10 KEGG pathways with raw pvalue < 0.05 from the epigenome-wide association study of cerebrospinal fluid-based (CSF) biomarkers of Alzheimer’s disease in the Emory Healthy Brain Study (EHBS, N=450).**

| **A. tTau** | | | | |
| --- | --- | --- | --- | --- |
| **KEGG pathway** | **Description** | **N** | **DE** | **P.DE** |
| hsa04114 | Oocyte meiosis | 117 | 15 | 3.97E-04 |
| hsa04714 | Thermogenesis | 214 | 19 | 0.002 |
| hsa04213 | Longevity regulating pathway - multiple species | 60 | 9 | 0.004 |
| hsa04371 | Apelin signaling pathway | 138 | 15 | 0.005 |
| hsa04662 | B cell receptor signaling pathway | 77 | 10 | 0.005 |
| hsa05218 | Melanoma | 70 | 9 | 0.01 |
| hsa04810 | Regulation of actin cytoskeleton | 218 | 20 | 0.013 |
| hsa00970 | Aminoacyl-tRNA biosynthesis | 43 | 6 | 0.014 |
| hsa05166 | Human T-cell leukemia virus 1 infection | 210 | 18 | 0.016 |
| hsa00360 | Phenylalanine metabolism | 14 | 3 | 0.019 |
| **B. pTau** | | | | |
| **KEGG pathway** | **Description** | **N** | **DE** | **P.DE** |
| hsa04114 | Oocyte meiosis | 117 | 14 | 0.001 |
| hsa05031 | Amphetamine addiction | 66 | 10 | 0.003 |
| hsa04151 | PI3K-Akt signaling pathway | 344 | 28 | 0.013 |
| hsa00640 | Propanoate metabolism | 32 | 5 | 0.015 |
| hsa00360 | Phenylalanine metabolism | 14 | 3 | 0.018 |
| hsa05166 | Human T-cell leukemia virus 1 infection | 210 | 18 | 0.02 |
| hsa04922 | Glucagon signaling pathway | 99 | 10 | 0.026 |
| hsa05203 | Viral carcinogenesis | 186 | 15 | 0.026 |
| hsa05167 | Kaposi sarcoma-associated herpesvirus infection | 188 | 15 | 0.03 |
| hsa04150 | mTOR signaling pathway | 152 | 14 | 0.03 |
| **C. Aβ42/tTau** | | | | |
| **KEGG pathway** | **Description** | **N** | **DE** | **P.DE** |
| hsa04392 | Hippo signaling pathway - multiple species | 29 | 6 | 0.007 |
| hsa00030 | Pentose phosphate pathway | 26 | 4 | 0.017 |
| hsa01200 | Carbon metabolism | 105 | 9 | 0.02 |
| hsa04512 | ECM-receptor interaction | 87 | 10 | 0.023 |
| hsa04928 | Parathyroid hormone synthesis, secretion and action | 113 | 12 | 0.03 |
| hsa04151 | PI3K-Akt signaling pathway | 344 | 25 | 0.036 |
| hsa01230 | Biosynthesis of amino acids | 67 | 6 | 0.041 |
| **D. Aβ42+/-** | | | | |
| **KEGG pathway** | **Description** | **N** | **DE** | **P.DE** |
| hsa00630 | Glyoxylate and dicarboxylate metabolism | 30 | 6 | 5.85E-04 |
| hsa01210 | 2-Oxocarboxylic acid metabolism | 31 | 6 | 8.30E-04 |
| hsa01230 | Biosynthesis of amino acids | 67 | 9 | 8.65E-04 |
| hsa05223 | Non-small cell lung cancer | 71 | 10 | 0.007 |
| hsa00290 | Valine, leucine and isoleucine biosynthesis | 4 | 2 | 0.01 |
| hsa00020 | Citrate cycle (TCA cycle) | 28 | 4 | 0.019 |
| hsa01200 | Carbon metabolism | 105 | 9 | 0.019 |
| hsa04152 | AMPK signaling pathway | 118 | 12 | 0.02 |
| hsa04068 | FoxO signaling pathway | 128 | 11 | 0.034 |
| hsa04730 | Long-term depression | 58 | 7 | 0.039 |
| **E. pTau+/-** | | | | |
| **KEGG pathway** | **Description** | **N** | **DE** | **P.DE** |
| hsa04921 | Oxytocin signaling pathway | 152 | 19 | 3.30E-04 |
| hsa04020 | Calcium signaling pathway | 242 | 25 | 3.99E-04 |
| hsa04750 | Inflammatory mediator regulation of TRP channels | 97 | 14 | 4.96E-04 |
| hsa04270 | Vascular smooth muscle contraction | 132 | 15 | 0.001 |
| hsa04720 | Long-term potentiation | 64 | 10 | 0.002 |
| hsa04022 | cGMP-PKG signaling pathway | 160 | 17 | 0.002 |
| hsa04724 | Glutamatergic synapse | 114 | 14 | 0.004 |
| hsa04925 | Aldosterone synthesis and secretion | 95 | 12 | 0.005 |
| hsa05208 | Chemical carcinogenesis - reactive oxygen species | 202 | 15 | 0.005 |
| hsa04929 | GnRH secretion | 62 | 9 | 0.006 |

**Table S10. First 10 KEGG pathways with raw pvalue < 0.05 from the epigenome-wide association study of cerebrospinal fluid-based (CSF) biomarkers of Alzheimer’s disease in ADNI (N=122).**

| **A. tTau** | | | | |
| --- | --- | --- | --- | --- |
| **KEGG pathway** | **Description** | **N** | **DE** | **P.DE** |
| hsa04015 | Rap1 signaling pathway | 207 | 20 | 0.008 |
| hsa05320 | Autoimmune thyroid disease | 43 | 4 | 0.024 |
| hsa00512 | Mucin type O-glycan biosynthesis | 35 | 5 | 0.03 |
| hsa00310 | Lysine degradation | 61 | 7 | 0.03 |
| hsa00982 | Drug metabolism - cytochrome P450 | 67 | 4 | 0.034 |
| hsa04940 | Type I diabetes mellitus | 38 | 4 | 0.037 |
| hsa05330 | Allograft rejection | 31 | 3 | 0.048 |
| **B. pTau** | | | | |
| **KEGG pathway** | **Description** | **N** | **DE** | **P.DE** |
| hsa04670 | Leukocyte transendothelial migration | 109 | 10 | 0.022 |
| hsa04928 | Parathyroid hormone synthesis, secretion and action | 113 | 11 | 0.048 |
| **C. Aβ42/tTau** | | | | |
| **KEGG pathway** | **Description** | **N** | **DE** | **P.DE** |
| hsa04928 | Parathyroid hormone synthesis, secretion and action | 113 | 17 | 1.33E-04 |
| hsa04916 | Melanogenesis | 101 | 13 | 0.001 |
| hsa00830 | Retinol metabolism | 65 | 6 | 0.001 |
| hsa04742 | Taste transduction | 83 | 8 | 0.006 |
| hsa00400 | Phenylalanine, tyrosine and tryptophan biosynthesis | 6 | 2 | 0.01 |
| hsa04022 | cGMP-PKG signaling pathway | 160 | 15 | 0.011 |
| hsa00250 | Alanine, aspartate and glutamate metabolism | 36 | 5 | 0.012 |
| hsa04062 | Chemokine signaling pathway | 189 | 15 | 0.012 |
| hsa04390 | Hippo signaling pathway | 155 | 15 | 0.013 |
| hsa04918 | Thyroid hormone synthesis | 73 | 8 | 0.021 |
| **D. Aβ42+/-** | | | | |
| **KEGG pathway** | **Description** | **N** | **DE** | **P.DE** |
| hsa04728 | Dopaminergic synapse | 128 | 13 | 0.014 |
| hsa04928 | Parathyroid hormone synthesis, secretion and action | 113 | 11 | 0.041 |
| hsa04022 | cGMP-PKG signaling pathway | 160 | 13 | 0.043 |
| hsa04927 | Cortisol synthesis and secretion | 63 | 7 | 0.05 |
| **E. pTau+/-** | | | | |
| **KEGG pathway** | **Description** | **N** | **DE** | **P.DE** |
| hsa05220 | Chronic myeloid leukemia | 74 | 11 | 0.003 |
| hsa04662 | B cell receptor signaling pathway | 78 | 10 | 0.005 |
| hsa05164 | Influenza A | 154 | 13 | 0.006 |
| hsa05161 | Hepatitis B | 153 | 15 | 0.006 |
| hsa05230 | Central carbon metabolism in cancer | 67 | 9 | 0.01 |
| hsa00051 | Fructose and mannose metabolism | 33 | 5 | 0.012 |
| hsa05210 | Colorectal cancer | 85 | 10 | 0.015 |
| hsa04977 | Vitamin digestion and absorption | 25 | 4 | 0.018 |
| hsa05163 | Human cytomegalovirus infection | 219 | 18 | 0.019 |
| hsa05219 | Bladder cancer | 40 | 6 | 0.02 |

**Table S11. First 10 KEGG pathways with raw pvalue < 0.05 from the epigenome-wide meta-analysis of cerebrospinal fluid-based (CSF) biomarkers of Alzheimer’s disease in in 450 cognitively normal individuals from the Emory Healthy Brain Study (EHBS) and 122 cognitively normal individuals from ADNI.**

| **A. tTau** | | | | |
| --- | --- | --- | --- | --- |
| **KEGG pathway** | **Description** | **N** | **DE** | **P.DE** |
| hsa04211 | Longevity regulating pathway | 88 | 12 | 0.003 |
| hsa04114 | Oocyte meiosis | 117 | 13 | 0.003 |
| hsa04213 | Longevity regulating pathway - multiple species | 60 | 9 | 0.004 |
| hsa04810 | Regulation of actin cytoskeleton | 218 | 20 | 0.014 |
| hsa00970 | Aminoacyl-tRNA biosynthesis | 43 | 6 | 0.014 |
| hsa04662 | B cell receptor signaling pathway | 77 | 9 | 0.016 |
| hsa00360 | Phenylalanine metabolism | 14 | 3 | 0.018 |
| hsa04714 | Thermogenesis | 214 | 16 | 0.02 |
| hsa05203 | Viral carcinogenesis | 186 | 15 | 0.023 |
| hsa04150 | mTOR signaling pathway | 152 | 14 | 0.026 |
| **B. pTau** | | | | |
| **KEGG pathway** | **Description** | **N** | **DE** | **P.DE** |
| hsa00640 | Propanoate metabolism | 32 | 7 | 4.96E-04 |
| hsa04390 | Hippo signaling pathway | 155 | 16 | 0.012 |
| hsa00360 | Phenylalanine metabolism | 14 | 3 | 0.017 |
| hsa03083 | Polycomb repressive complex | 75 | 9 | 0.017 |
| hsa04114 | Oocyte meiosis | 117 | 11 | 0.021 |
| hsa00350 | Tyrosine metabolism | 34 | 4 | 0.031 |
| hsa04213 | Longevity regulating pathway - multiple species | 60 | 7 | 0.041 |
| **C. Aβ42/tTau** | | | | |
| **KEGG pathway** | **Description** | **N** | **DE** | **P.DE** |
| hsa04730 | Long-term depression | 58 | 11 | 2.72E-04 |
| hsa04928 | Parathyroid hormone synthesis, secretion and action | 113 | 14 | 0.005 |
| hsa04392 | Hippo signaling pathway - multiple species | 29 | 6 | 0.006 |
| hsa04270 | Vascular smooth muscle contraction | 132 | 12 | 0.026 |
| hsa01200 | Carbon metabolism | 105 | 8 | 0.043 |
| **D. Aβ42+/-** | | | | |
| **KEGG pathway** | **Description** | **N** | **DE** | **P.DE** |
| hsa04722 | Neurotrophin signaling pathway | 114 | 12 | 0.011 |
| hsa04744 | Phototransduction | 27 | 4 | 0.013 |
| hsa03015 | mRNA surveillance pathway | 85 | 8 | 0.017 |
| hsa04261 | Adrenergic signaling in cardiomyocytes | 148 | 14 | 0.019 |
| hsa04114 | Oocyte meiosis | 117 | 10 | 0.024 |
| hsa01200 | Carbon metabolism | 105 | 8 | 0.035 |
| hsa05214 | Glioma | 74 | 8 | 0.039 |
| hsa00130 | Ubiquinone and other terpenoid-quinone biosynthesis | 11 | 2 | 0.042 |
| hsa05225 | Hepatocellular carcinoma | 166 | 13 | 0.048 |
| **E. pTau+/-** | | | | |
| **KEGG pathway** | **Description** | **N** | **DE** | **P.DE** |
| hsa04724 | Glutamatergic synapse | 114 | 14 | 0.002 |
| hsa04270 | Vascular smooth muscle contraction | 132 | 12 | 0.011 |
| hsa04929 | GnRH secretion | 62 | 8 | 0.012 |
| hsa04720 | Long-term potentiation | 64 | 8 | 0.012 |
| hsa04068 | FoxO signaling pathway | 128 | 11 | 0.013 |
| hsa04722 | Neurotrophin signaling pathway | 114 | 11 | 0.014 |
| hsa04936 | Alcoholic liver disease | 136 | 10 | 0.017 |
| hsa05170 | Human immunodeficiency virus 1 infection | 200 | 14 | 0.019 |
| hsa05169 | Epstein-Barr virus infection | 188 | 13 | 0.02 |
| hsa05220 | Chronic myeloid leukemia | 74 | 8 | 0.022 |
